# The impact of the COVID-19 pandemic on the provision & utilisation of primary health care services in Goma, Democratic Republic of the Congo, Kambia district, Sierra Leone & Masaka district, Uganda

**DOI:** 10.1101/2022.04.28.22274416

**Authors:** K Kasonia, D Tindanbil, J Kitonsa, K Baisley, F Zalwango, L Enria, A Mansaray, M James, Y Nije, D Tetsa Tata, B J Lawal, A Drammeh, B Lowe, D Mukadi-Bamuleka, S Mounier-Jack, F Nakiyimba, P Obady, J Muhavi, J S Bangura, B Greenwood, M Samai, B Leigh, D Watson-Jones, H Kavunga-Membo, E Ruzagira, K E. Gallagher

## Abstract

**Introduction:** This study aimed to determine whether the COVID-19 pandemic had an impact on the number of people seen at public facilities in Uganda, the Democratic Republic of the Congo (DRC) and Sierra Leone for essential primary healthcare services.

**Methods:** The number of weekly consultations for antenatal care (ANC), outpatient (OPD), expanded programme on immunisations (EPI), family planning (FP) services and HIV, for the period of January 2018-December 2020, were collected from 25 primary healthcare facilities in Masaka district, Uganda, 21 health centres in Goma, DRC, and 29 facilities in Kambia district, Sierra Leone. Negative binomial regression models accounting for facility level clustering and season were used to analyse changes in activity levels between 2018, 2019 and 2020.

**Results:** We found no evidence that the COVID-19 pandemic affected the number of OPD, EPI or ANC consultations in Goma. Family planning consultations were 17% lower in March-July 2020 compared to 2019, but this recovered by December 2020. New diagnoses of HIV were 34% lower throughout 2020 compared to 2019. Compared to the same periods in 2019, facilities in Sierra Leone had 18-29% fewer OPD consultations throughout 2020, and 27% fewer DTP3 doses in March-July 2020, but this had recovered by Jul-Dec. There was no evidence of differences in other services. In Uganda there were 20-35% fewer under-5 OPD consultations, 21-66% fewer MCV1 doses, and 48-51% fewer new diagnoses of HIV, throughout 2020, compared to 2019. There was no difference in the number of HPV doses delivered in 2020 compared to 2019.

**Conclusions:** The level of disruption appeared to correlate with the strength of lockdown measures in the different settings and community attitudes towards the risk posed by COVID-19. Mitigation strategies such as health communications campaigns and outreach services proved important to limit the impact of lockdowns on primary healthcare services.

**Key messages:** *What is already known on this topic:* The COVID-19 pandemic and the response measures put in place caused disruption to the provision and utilisation of primary healthcare services worldwide.

*What this study adds:* We document that the COVID-19 pandemic had a varied impact on different services in three distinct settings on the African continent. The extent that the pandemic impacted services correlated with the stringency of the lockdowns, community perceptions of the level of danger posed by the pandemic and communities’ prior exposure to Ebola epidemics and concomitant response measures.

*How this study might affect research, practice, or policy:* strategies such as communication campaigns and outreach services limited the impact of lockdowns on essential services and would be valuable strategies to implement in future epidemics.

## Introduction

The World Health Organization (WHO) pulse surveys reported that the COVID-19 pandemic affected the provision and utilization of essential primary healthcare services in >90% of countries worldwide. In the first survey, conducted between May and July 2020, 25 essential services were assessed in 105 countries. Nearly all countries reported either partial (5%-50%) or severe (>50%) change in service provision or use. Low and lower-middle income countries were more affected than countries in higher income brackets(1). In the second survey, conducted between January and March 2021, 94% of the 135 countries that took part in the survey reported residual service disruption(2). In response to the pandemic, various containment measures such as social distancing, lockdowns, curfews, closure of schools and bans on gatherings were instituted across the globe(3). Fewer transport options and less disposable income during lockdowns, alongside fears and misconceptions around the risk and feasibility of accessing services could have reduced utilisation of primary care. Healthcare resources, including staff, facilities, consumables, treatments, personal protective equipment, were re-prioritised to fight the disease, social distancing measures were put in place in facilities, and many of the workforce fell sick, reducing the capacity to provide essential health services in many settings(1).

Literature reviews have already documented that COVID-19 had a considerable impact on primary healthcare at both the service and patient level(4, 5); however, almost all studies are from Europe or the USA. The UK reported a 20% reduction in measles vaccinations, three weeks after social distancing measures were announced(6). In the USA, the timing of the pandemic correlated with a significant decrease in women arriving at a large, regional referral hospital with a contraception plan(7). A meta-analysis of 14 studies from countries in Europe, Asia and one in Africa reported a 33% increase in the rate of stillbirth during lockdown (95% CI 1.04,1.69)(5). There is a paucity of information about the impact of the pandemic on the provision and utilization of primary healthcare services in low-and middle-income countries (LMICs), especially in Africa. A study in Rwanda documented a significant decrease in utilization of antenatal care, facility-based deliveries, post-natal care and vaccinations when comparing April-May 2020 with April-May 2019(8). In Uganda, a 75% reduction in HIV testing and initiation of antiretroviral treatment was reported in the first three weeks of April 2020 compared to the weekly average for the period from Jan-Mar 2020(9).

The first case of SARS-CoV-2 infection in the Democratic Republic of the Congo (DRC) was identified on 10^th^ March 2020. From the 18^th^ March onwards, in response to the pandemic, the government put in place movement restrictions, closure of public spaces, limitations on gatherings and compulsory wearing of masks in public (10). The first case of SARS-CoV-2 infection in Sierra Leone was recorded on 30^th^ March 2020(11). The President of the Republic of Sierra Leone declared a public health emergency for twelve months and imposed a dusk to dawn curfew, movement restrictions, bans on public gatherings, school closures, and compulsory wearing of masks. The first case of SARS-CoV-2 infection was confirmed in Uganda on 21^st^ March 2020(12). Subsequently, the Ministry of Health imposed movement restrictions, closure of public spaces and schools, gatherings were limited, and masks were made compulsory in public. All these measures may have affected effective provision and utilization of primary health services in these countries.

We aimed to determine in what ways and to what extent the pandemic impacted provision and utilization of primary healthcare services in 2020 in three distinct settings with different documented burdens of COVID-19 and different lockdown measures. We focus on the number of people seen at healthcare facilities for antenatal care, under-5 outpatient services, routine immunisations, family planning and HIV treatment services, using routine health registration data. Qualitative data were collected and inform the discussion of the data presented in this paper but are reported separately. A French translation of this manuscript is provided in supplementary files.

## Methods

### Study setting & health facility selection

The study was conducted in three areas, Goma, in the Democratic Republic of the Congo, Kambia District, north-western Sierra Leone, and Masaka District, south-western Uganda (Table 1, 2). District/ regional authorities were approached for approval to conduct the study and complete lists of health facilities in each area were compiled. Private health facilities were excluded from selection, as the aim of the project was to inform the provision of public health services. A selection of health centres was made to include all 25 available government health centres in Masaka, all 21 accessible health centres in urban Goma; and a random number generator was used to select a representative selection of 29 health facilities in Kambia, proportional to the total number of health posts and health centres in the district.

**Table 1.** The study setting and selected health centres.

| Country | Selected region/district | Description and total population | Total public health facilities in the area | Selected facilities for quantitative data collection | Selected facilities for qualitative interviews |
| --- | --- | --- | --- | --- | --- |
| DRC | Goma | Urban and suburban; estimated population: 600,000- 1 million | 39 (26 health centres, 13 tertiary care hospitals) | 21 (21 health centres<sup>1</sup>) | 12 |
| Sierra Leone | Kambia | Suburban and rural; estimated population: 350,000 | 68 (55 health posts, 15 health centres, 1 hospital) | 29 (22 health posts, 6 health centres, 1 hospital with a primary health care dept.<sup>2</sup>) | 15 |
| Uganda | Masaka | Rural; estimated population: 307,000 | 26 (25 health facilities, 1 referral Hospital) | 25 (14 level II, 9 level III, 2 level IV health centres<sup>3</sup>) | 15 |

<sup>1</sup> In Goma, health centres provide primary healthcare to the urban population; 21 of the 26 public health centres were selected due to security and logistical constraints during data collection.
<sup>2</sup> In Kambia, health posts, community health centres and some hospitals provide primary care to the population. Health posts (community or maternal child health posts) are small and provide services in remote rural settings; community health centres are larger, based in central locations within each chiefdom and can have laboratory and more consistent cold chain services. Kambia district hospital was included as the dominant public primary health care provider within Kambia town.
<sup>3</sup> In Masaka, level II health facilities provide outpatient services to the surrounding parish, Level III facilities are slightly larger sub-county facilities which may have maternity wards, and level IV facilities have inpatient and outpatient services.

**Table 2.** Definition of periods of analysis based on the COVID pandemic and lockdown measures.

| Area | Period | Approx. dates in the period (2018-20)<sup>1</sup> | N (weeks/year) | Description of lockdown measures in place in 2020 | Mean lockdown stringency index for the period(3) | National number of reported COVID-19 cases(15) | National number of cases per 1 million population |
| --- | --- | --- | --- | --- | --- | --- | --- |
| Goma | 0 | 1<sup>st</sup> January – 22<sup>nd</sup> March | 12 | Pre-COVID | 0 |  |  |
|  | 1 | 23<sup>rd</sup> March – 19<sup>th</sup> July | 17 | First wave of reported cases peaks. State of emergency (declared 18<sup>th</sup> March), stay at home order for 14 days, schools closed. | 80 | 8443 | 94.3 |
|  | 2 | 20<sup>th</sup> July – 18<sup>th</sup> October | 12 | Limitations on gatherings lifted, public places reopened. | 49 | 2557 | 28.6 |
|  | 3 | 19<sup>th</sup> October – 27<sup>th</sup> December | 10 | Cases begin to rise in second wave. Low stringency lockdown, restrictions start to be implemented again mid-December. | 26 | 5839 | 65.2 |
| Kambia | 0 | 1<sup>st</sup> January – 15<sup>th</sup> March | 11 | Pre-COVID | 0 |  |  |
|  | 1 | 16<sup>th</sup> March – 19 July | 18 | First wave of reported infections. Gatherings of over 100 banned, state of emergency (declared 31 March). | 60 | 1711 | 214.5 |
|  | 2 | 20<sup>th</sup> July – 27<sup>t</sup> December | 22 | Low number of cases and low stringency lockdown measures. | 33 | 849 | 106.4 |
| Masaka | 0 | 1<sup>st</sup> January – 15<sup>th</sup> March | 11 | Pre-COVID | 0 |  |  |
|  | 1 | 16<sup>th</sup> March – 20<sup>th</sup> September | 26 | Gatherings restricted, schools closed, nationwide lockdown, transport restrictions. | 83 | 6287 | 137.5 |
|  | 2 | 21<sup>st</sup> September – 27<sup>th</sup> December | 13 | Some public places re-opened, curfew remained in place. | 59 | 27524 | 601.7 |

<sup>1</sup> Each year was classified into 52 weeks and data was analysed by week, the dates included in each period varied slightly from year to year, dates provided in the table are those corresponding to the selected weeks in 2020, which defined the periods of analysis. The periods of analysis were selected based on COVID-19 case numbers and the implementation of lockdown measures.

### Data collection

Trained staff visited each of the selected facilities and collected general information from the head of each health facility, detailing: the facility location, the number of staff members, the services provided and estimated catchment population, and any other known service disruptions over the study period. Project staff then tallied weekly counts of the number of consultations in the registration book for each service from January 2018 to December 2020. Data were entered directly onto an electronic REDcap database on computer tablets(13, 14). Data were collected on the weekly number of under-5 outpatient department (OPD) consultations, first antenatal care (ANC) visits, third doses of the diphtheria-tetanus-pertussis (DTP3) vaccine delivered, first doses of measles-containing vaccine (MCV1) delivered, first doses of human papillomavirus vaccine (HPV1) delivered, family planning (FP) consultations, new HIV diagnoses and HIV care visits (including for ART replenishment). If data were available on the probable diagnosis at the OPD visit, the numbers of children diagnosed with respiratory diseases and probable malaria were noted, and this was further broken down by whether the child was referred for admission or treated as an outpatient.

### Sample size estimation and statistical analysis

The Health Management Information System (HMIS) data for Masaka District, Uganda, was used to estimate the sample size required for the study to detect a relative change of 30% when comparing a single week in 2020 with the same week the year before. Assuming an average of 157 OPD visits per week, with a standard deviation of 73, a sample of at least 20 facilities per period would enable the study to detect a relative change of 30% with 80% power at the 5% significance level.

The period of analysis was from the 1^st^ January 2018 to the 27^th^ December 2020. Data availability was assessed as the number of facilities with data for each week of analysis, this was plotted over time for each area, by service. The average number of consultations per facility with data, per week, was calculated and plotted against time for each service in each area. A pre-pandemic period was defined as January-March 2020 (Table 2). Distinct periods of the pandemic were defined using the reported ‘waves’ of COVID-19 using Ministry of Health (MOH) data in each area, reported lockdown measures, and the estimated ‘stringency’ of the lockdown in each setting(3) (Table 2). For each defined period of analysis, negative binomial regression was used to estimate the relative change in count, comparing 2018, 2019 and 2020, using robust standard errors to adjust for autocorrelation and controlling for ‘month’ as a season parameter. Clinic level random effects were included to account for between clinic differences and within-clinic clustering of data. Climate data from the nearest available weather station were downloaded and assessed in the model as a potential confounder.

### Patient & public involvement

Local health authorities were engaged in the conceptualisation and implementation of the study; the public were not involved in the conceptualisation or design of the study. This study was approved by the Comite National d’Ethique de la Sante (CNES) of the DRC, the Sierra Leone Ethics and Scientific Review committee, the Uganda Virus Research Institute Research Ethics Committee, the Uganda National Council for Science and Technology, the London School of Hygiene and Tropical Medicine Ethics Committee and local health authorities in each area. No informed consent was required for this study as individual level data were not collected.

## Results

The three settings were distinct in health service organisation and context. Facilities in Goma were relatively large, with an average catchment population of 30,000 and on average 12 registered/trainee nurses per facility in 2020. Masaka facilities served an average of 11,000 people and had an average of 4 registered/trainee nurses per facility in 2020. In Kambia, primary healthcare facilities were generally small, serving 5,000 people and were staffed by on average just 1 registered/ trainee nurse alongside supporting staff (community health volunteers, midwives etc; Table 3).

**Table 3.** Catchment populations and staffing over time in the selected facilities.

| Facility characteristics | Goma |  |  | Kambia |  |  | Masaka |  |  |
| --- | --- | --- | --- | --- | --- | --- | --- | --- | --- |
|  | 2018 | 2019 | 2020 | 2018 | 2019 | 2020 | 2018 | 2019 | 2020 |
| **Catchment population** |  |  |  |  |  |  |  |  |  |
| Mean per facility (s.d.) | 28,048 | 29,137 | 30,011 | 4,493 | 5,113 | 5,078 | 10,076 | 9,622 | 11,399 |
| Range per facility | 9,820-52,733 | 10,115 – 54,315 | 10,418 - 55,944 | 2,759-7,400 | 1,168 – 17,724 | 1,259-19,318 | 1,652-25,877 | 1,686-26,415 | 1,721-50,381 |
| Number of facilities with data | 21 | 21 | 21 | 15 | 18 | 21 | 24 | 23 | 24 |
| **Staff** |  |  |  |  |  |  |  |  |  |
| Mean number of **registered/ trainee nurses** per facility (range) | 10.5 (7-18) | 11.2 (4-21) | 11.9 (2-21) | ~ | ~ | 0.7 (0-3) | ~ | ~ | 3.8 (0-13) |
| Number of facilities with data | 14 | 15 | 21 | 0 | 0 | 29 | 0 | 0 | 25 |

~: missing data; s.d.: standard deviation

### Goma, DRC

Data were available from all 21 facilities in 2019 and 2020 for all services; some facilities were missing data for some services in 2018 (**Supplementary figure 1**). Mean counts of service activity per facility per week were highly variable across all periods and all years (**Supplementary figure 2**). In the calendar period of January-March, point estimates indicate that service activity may have been increasing across the years (2018, 2019, 2020) for many services (**Supplementary table 1**). There were significantly higher numbers of consultations for OPD (relative risk (RR) 1.38 (95% confidence interval (CI) 1.09-1.74)), DTP3 (RR 1.21 (95%CI 1.07-1.36) and ANC services (RR 1.26 (95%CI 1.07-1.49) in 2020 compared to 2019. There was no evidence of a difference between 2020 and 2019 in the number of consultations for other services in this period.

In period 1 of the pandemic (March-July), there was no evidence of a difference in the number of OPD consultations in 2020 compared to 2019 and this was sustained in periods 2 (July-October) and 3 (October-December; **Figure 1**). There was some indication that visits to OPD for respiratory complaints had decreased but malaria OPD visits had increased in period 1 of lockdown, compared to similar periods in 2018 and 2019, but this difference disappeared in periods 2 and 3. The number of DTP3 doses delivered in period 1 of the pandemic was 20% higher in 2020 compared to similar calendar months in 2019 (95%CI 7-36), but there was no evidence of a difference between 2020 and 2019 in periods 2 and 3. The point estimates for the difference in the number of MCV1 doses delivered indicated a 12-18% higher number of doses in 2020 compared to 2019; this was only statistically significant in period 3 of the pandemic. The number of first ANC visits in period 1 and 3 of the pandemic were 17% (95%CI 4-32%) and 19% (95%CI 2-38) higher than in the same periods in 2019; there was no evidence of a difference in period 2. There was no evidence of a difference in TT doses delivered, relative to the number delivered in 2019, in any of the calendar periods. There was evidence of 17% fewer FP consultations in period 1 of the pandemic (95%CI 2-31), but this difference disappeared in periods 2 and 3. The number of new diagnoses of HIV were 34% lower in both period 1 (95%CI 20-47) and period 2 (95%CI 1-56) of lockdown compared to the same periods the year before, but this effect disappeared in period 3. There was no evidence of a change in repeat ART replenishment visits, in any of the periods (**Figure 1**).

**Figure 1.**
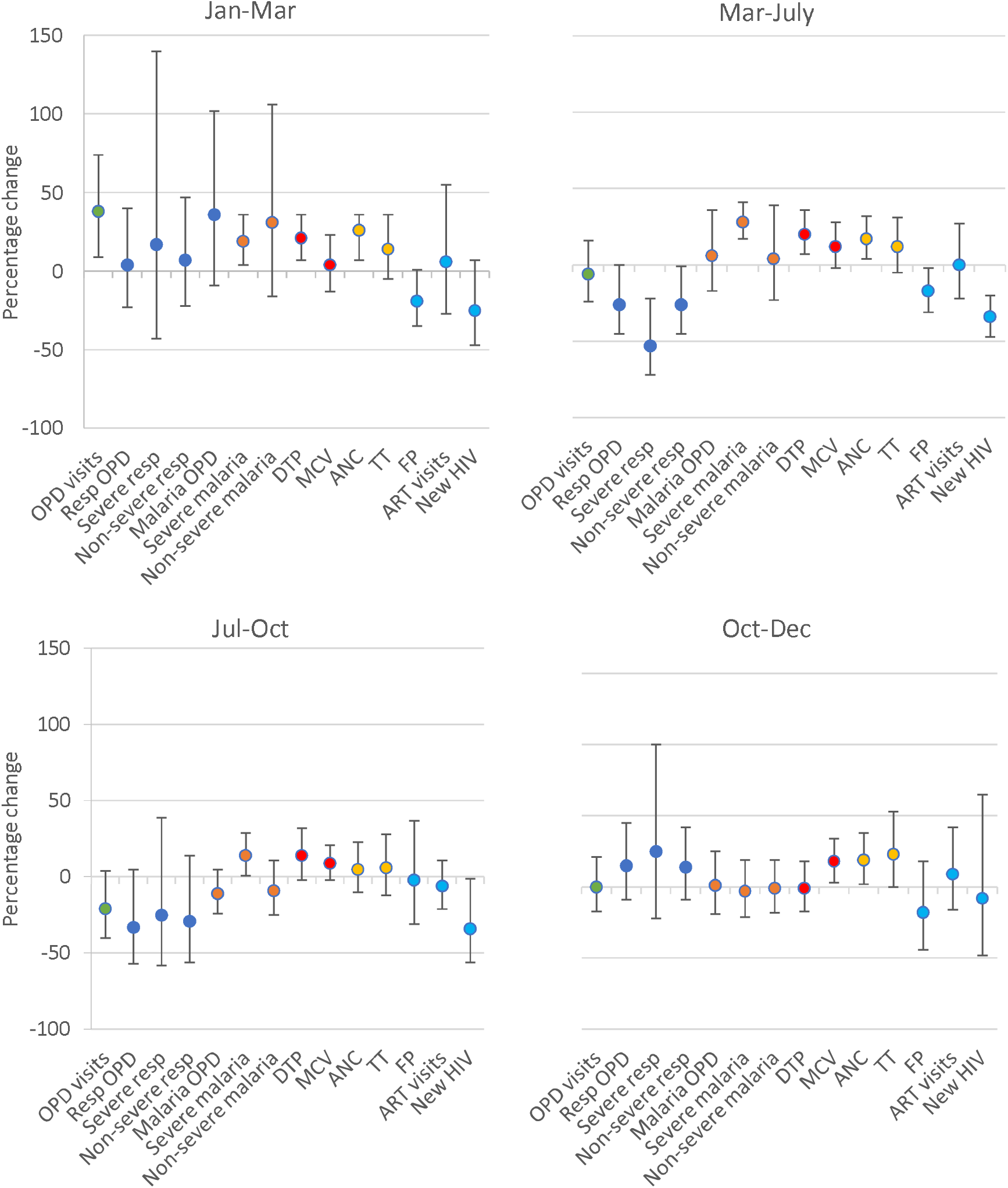
The percentage change in activity levels in 2020 compared to 2019, for each outcome, in each period, in Goma^1^. ^1^ The percentage change was calculated as (1-RR_2020/2019_)*100; the RRs for 2018 vs 2019 and 2019 vs 2020 are included in Supplementary Table 1

### Kambia, Sierra Leone

Data were available from 25-29 facilities for all of the major service groups except HIV services (**Supplementary figure 3**); only 5-6 facilities had data on HIV services for 2018-2020. Data on whether outpatients were referred to a higher-level facility or treated as outpatients were incomplete and not analysable. Mean counts of service activity per facility per week were highly variable across all periods and all years (**Supplementary figure 4**). In the pre-pandemic period (Jan-March), there was no evidence of a difference in the level of activity for any of the services across any of the three years 2018, 2019 and 2020 (**Supplementary table 2, Figure 2**).

**Figure 2.**
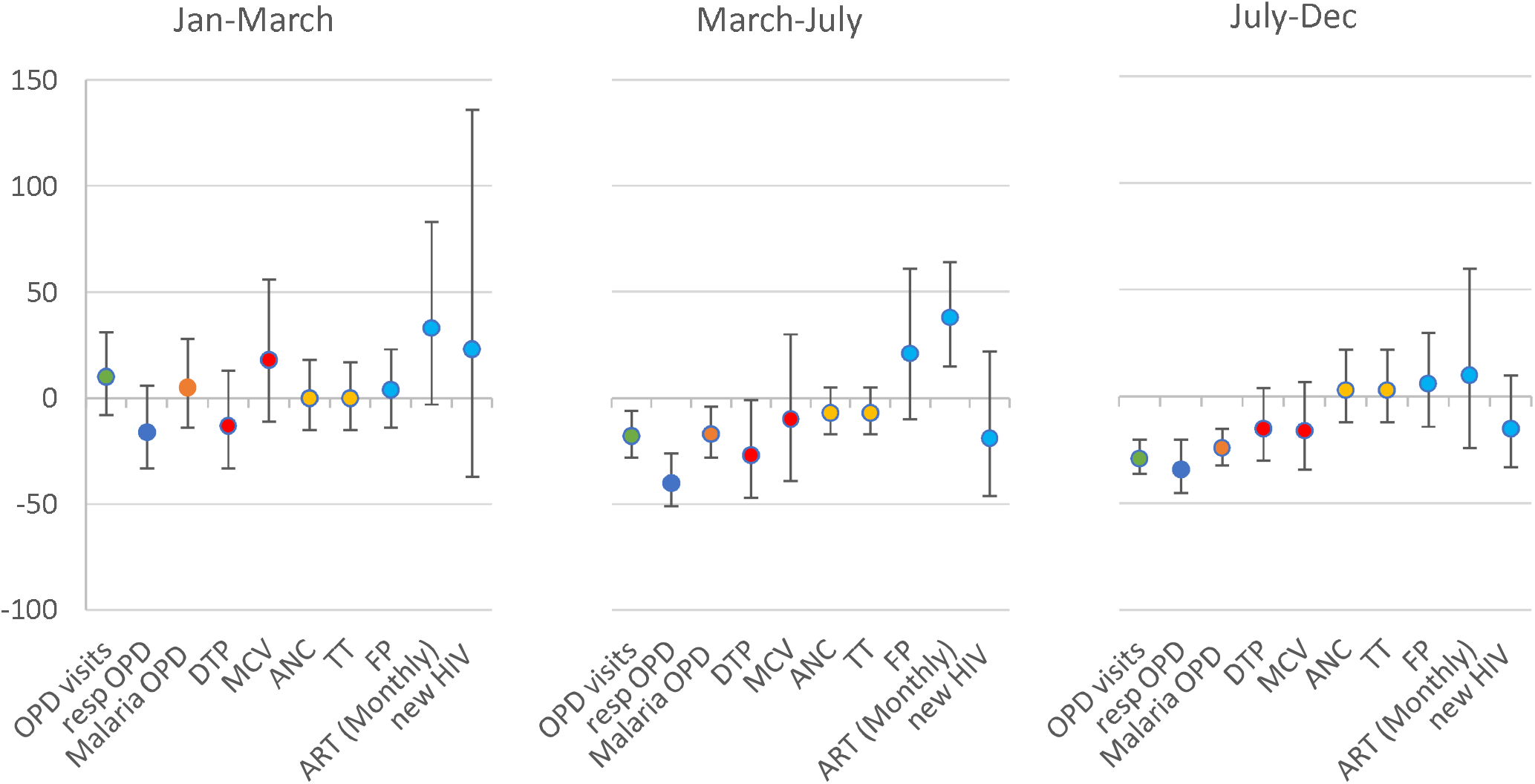
The percentage change in activity levels in 2020 compared to 2019, for each outcome, in each period, in Kambia, Sierra Leone^1^. ^1^ The percentage change was calculated as (1-RR_2020/2019_)*100; the RRs for 2018 vs 2019 and 2019 vs 2020 are included in Supplementary Table 2

In period 1 of the pandemic in Sierra Leone (March-July), there were 18% fewer OPD consultations, compared to similar periods in 2019; this reduction was sustained in period 2 (July-December) where there were 29% fewer OPD consultations (95%CI 20-36%) compared to the same period in 2019. When broken down by diagnosis, there were fewer respiratory and malaria OPD visits. There were 27% fewer DTP3 doses delivered in period 1 in 2020 compared to 2019 (95%CI 1-47%); but this difference disappeared in period 2. There was no evidence of a difference in the number of MCV1 doses delivered, ANC consultations, FP consultations or new HIV diagnoses in 2020 compared to 2019 in any of the periods of analysis, although CI are wide. In period 1 of the pandemic, there were 38% more repeat ART replenishment visits in 2020 compared to 2019, but this difference disappeared in period 2.

### Masaka, Uganda

Data were available from 20-25 facilities in 2019 and 2020 for most services; some facilities were missing data for some services in 2018 (**Supplementary figure 5**). HIV service data were only available from 12 facilities in 2020, compared to 15-16 facilities in 2019. Mean counts of service activity per facility per week were highly variable across all periods and all years (**Supplementary figure 6**). In the pre-pandemic period (Jan-March), there was no evidence of a difference in the level of activity for any of the services across any of the three years 2018, 2019 and 2020 (**Supplementary table 3, Figure 3**).

**Figure 3.**
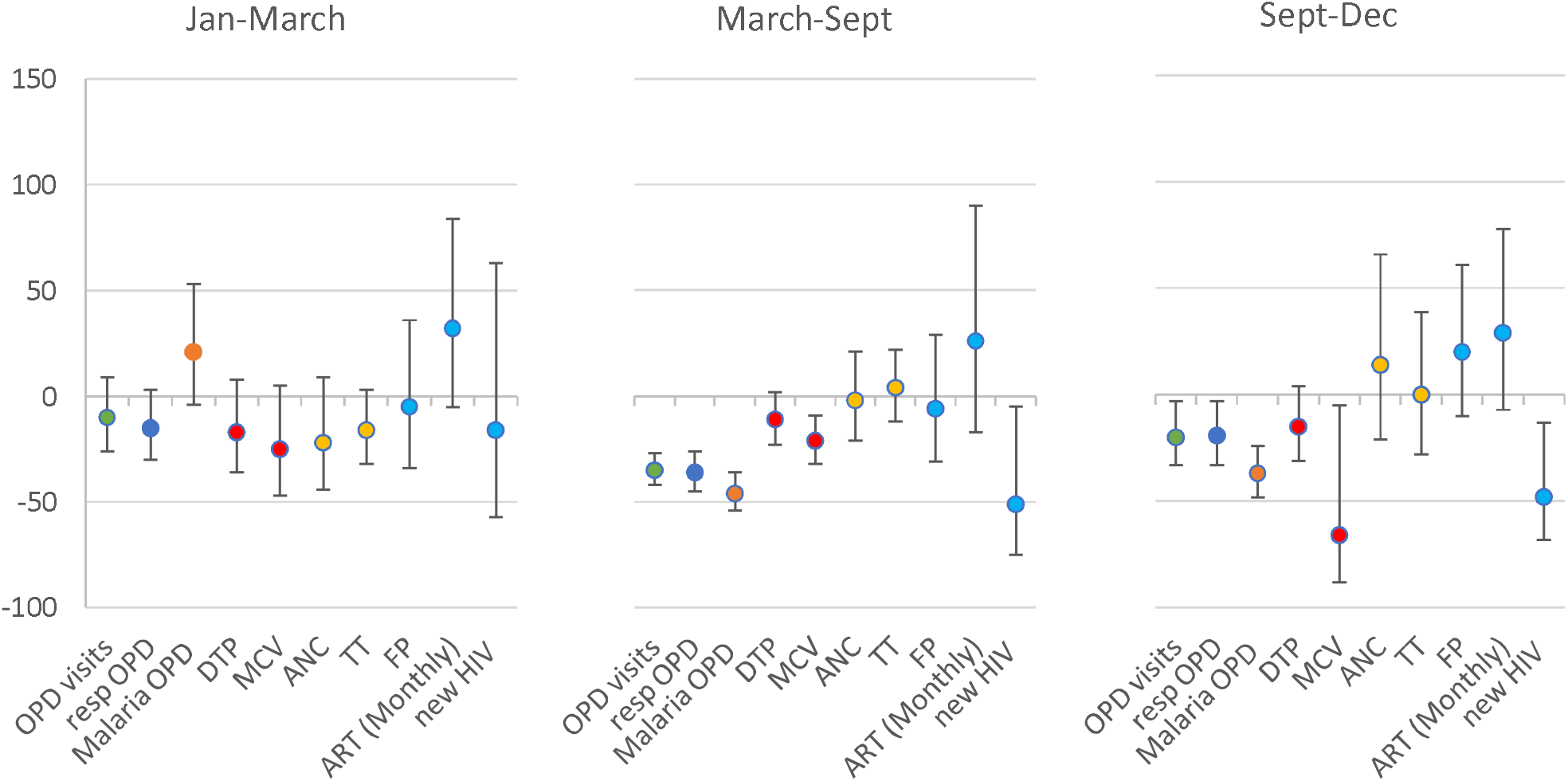
The percentage change in activity levels in 2020 compared to 2019, for each outcome, in each period, in Masaka, Uganda^1^. ^1^ The percentage change was calculated as (1-RR_2020/2019_)*100; the RRs for 2018 vs 2019 and 2019 vs 2020 are included in Supplementary Table 3

In period 1 of the pandemic in Uganda (March-September), there were 35% fewer OPD visits (95%CI 27-42) in 2020 compared to 2019, and this was sustained into period 2 (September-December) with 20% fewer OPD visits (95%CI 3-33; **Figure 3**) in 2020 compared to 2019. In period 1 and 2 of the pandemic, point estimates indicate 11-15% fewer DTP3 doses delivered in 2020 compared to 2019, although confidence intervals cross the null. There were 21% fewer MCV1 doses delivered in period 1 of the pandemic in 2020 compared to 2019 (95%CI 9-32), and this difference increased in period 2, with 66% fewer MCV1 doses than in the previous year (95%CI 5-88). There was no evidence of a difference in the number of consultations for ANC, FP or ART replenishment when comparing 2020 and 2019 service activity in any of the periods of analysis. The number of new HIV diagnoses in 2020 was half that of 2019, in both period 1 (51% reduction 95%CI 5-75) and period 2 (48% reduction 95%CI 13-68) of the pandemic.

Uganda was the only setting to deliver HPV vaccine in the study period. There were 77% fewer HPV vaccine doses delivered to girls aged 10 in period 1 of lockdown (March-September 2020), compared to 2019. This had bounced back with a substantial increase in doses in period 2 of lockdown. Overall, the number of first doses of HPV vaccine delivered in each year was similar.

## Discussion

In this analysis of weekly service activity levels across health facilities in Goma (DRC), Masaka (Uganda) and Kambia (Sierra Leone), we observed some reductions in primary care utilization/ provision during the lockdowns in 2020. The change in activity levels differed across services and settings (**Table 4**).

**Table 4.** Summary of percentage change in activity comparing 2020 with 2019 activity levels, for each period of lockdown by area.

| Area | Goma<sup>1</sup> |  |  | Kambia<sup>2</sup> |  | Masaka<sup>3</sup> |  |
| --- | --- | --- | --- | --- | --- | --- | --- |
| Service | Period 1 | Period 2 | Period 3 | Period 1 | Period 2 | Period 1 | Period 2 |
| OPD | -6% (-24, +16) | -21% (-40, +4) | 0 (-17, 21) | -18% (-6, -28) | -29% (-20, -36) | -35% (-27, -42) | -20% (-33, -3) |
| DTP | +20 (+7, +36) | +14% (-2, +32) | -1 (-17, 18) | -27% (-1, -47) | -15% (-30, +4) | -11% ((+2, -42) | -15% (-31, +4) |
| MCV | +12% (-2, +28) | +9% (-2, +21) | +18 (3, 34) | -10% (-39, +30) | -16% (-34, +7) | -21% (-9, -32) | -66% (-88, -5) |
| ANC | +17 (+4, +32) | +5% (-10, +23) | +19 (2, 38) | -7% (-17, +5) | +3% (-12, +22) | -2 (+21, -21) | +14, (-21, +66) |
| FP | -17% (-2, -31) | -2% (-31, +37) | -18 (-44, 18) | +21% (-10, +61) | +6% (-14, +30) | -6 (+29, -31) | +20 (-10, +61) |
| ART | 0% (-22, +27) | -6% (-21, +11) | +9 (-16, 42) | +38% (+15, +64) | +10% (-24, +60) | +26% (-17, +90) | +29 (-7, +78) |
| new HIV | -34% (-20, -47) | -34% (-56, -1) | -8 (-48, 65) | -19% (-46, +22) | -15% (-33, +10) | -51% (-5, -75) | -48 (-68, -13) |
| HPV dose 1 | ~ | ~ | ~ | ~ | ~ | -77% (-43, -81) | +84% (-9, +372) |

<sup>1</sup> In Goma, Period 1: 23 Mar – 19 Jul 2020, Period 2: 20 Jul – 18 Oct 2020; Period 3: 19 Oct – 27 Dec.
<sup>2</sup> In Kambia, Period 1: 16 Mar – 19 Jul 2020, Period 2: 20 July – 27 Dec 2020.
<sup>3</sup> In Masaka, Period 1: 16 Mar – 20 Sept 2020, Period 2: 21 Sept – 27 Dec 2020.

**In Goma, DRC**, we observed little to no difference in service activity levels in 2020 compared to 2019 and 2018. There were a higher number of vaccinations delivered in the pandemic periods of 2020 compared with 2019. We observed fewer family planning consultations, which is likely to reflect a true unmet need as the International Red Cross had discontinued funding family planning in Goma in 2020 (pers. Comm. P Obady). The reduction in HIV diagnoses reflects a reported stockout of diagnostic tests for HIV during the COVID 19 pandemic. The limited impact of the pandemic on routine primary healthcare utilisation or provision was surprising considering that qualitative data provided by healthcare workers described reduced staffing and disrupted services during this period (M. James et al. *manuscript in preparation*). Qualitative data from community members suggested some distrust among community members and fears that if they attended facilities, they might be forcibly quarantined, infected with COVID-19 or given the vaccine (F. Zalwango et al. *manuscript in preparation*). However, our findings may be explained by other qualitative data reflecting the perceived low risk of COVID-19 disease within a health system which was only just emerging from a recent Ebola epidemic(16), and past expertise in responding to outbreaks of public health concern. Temperature screening and infection prevention and control measures had been in place in primary health facilities for some time to screen for Ebola and the community were familiar with these measures. Among health management, COVID-19 was perceived from the beginning as ‘not as bad as Ebola’. However, crisis response teams conducted substantial community engagement, delivering messages on preventative measures and the availability of health services. Cholera vaccination campaigns, religious leaders and women’s associations were all targeted with educational messages about COVID-19 and the services available at the health centre. Additionally, the lockdown within Goma was stringent to begin with but eased relatively quickly and was variably adhered to; crucially, transport options remained available for people to get to health centres.

**In Kambia, Sierra Leone**, we observed some reductions in OPD and DTP3 activity levels during the pandemic, with approximately 20% fewer OPD consultations and 30% fewer DTP3 doses delivered in Mar-July 2020 compared to the same period in 2019. Qualitative data from community members described fears of getting infected at the health facility, forced vaccination and vaccine side-effects and fears of being diagnosed with COVID-19 and the potential consequences (including quarantine). Health care workers also reported reduced staffing at facilities. However, experiences during the Ebola epidemic of 2014-16 may have mitigated some of the impact of lockdown measures. Social mobilisation campaigns were held to encourage the population to access healthcare if they needed it for COVID testing, vaccines and routine services. We observed an increase in HIV ART replenishment visits, potentially explained by health communication messages at the time that encouraged people living with HIV to attend the facility to collect a 3-month supply of ART due to uncertainties around service continuation at the beginning of the pandemic.

**In Masaka, Uganda**, we observed substantial reductions in service activity for several services including OPD, DTP3, MCV1, HIV care and HPV vaccination. The lockdown enforced during the pandemic was stringent and prolonged with reduced transport options and a 7.00pm to 06:30am curfew. Stresses and strains on the health service during the pandemic were substantial. Healthcare workers were out of station with sickness and quarantine and one reportedly died. Although the usual outreach for integrated mother and child health services continued and potentially mitigated the impact of the lockdown on FP, ANC and infant immunisation service uptake, there was initially no additional budget for additional outreach services e.g. for OPD and HIV care, as it had not been anticipated that these would be needed. Our findings are supported by another study documenting a 77% decrease in new HIV diagnoses in the first weeks of April 2020 compared to January-March 2020(9); additional to this previous paper, we found that that this decrease was sustained throughout 2020.

HPV vaccine is usually delivered in Uganda via school-based outreach programmes; although schools remained closed throughout 2020, the MOH supplied a specific grant to support HPV vaccine delivery through community outreach, integrated with child health days, later in 2020. This mitigation strategy worked and by the end of the year, the number of girls who had received their first dose of HPV was no different to previous years. MCV1 was given at these health days but uptake may have suffered from lack of mobilisation or awareness as provision was substantially lower in 2020 compared to 2019. Measles catch up campaigns would be recommended in this setting to avoid outbreaks.

The data we report aligns with a worldwide analysis which documented a 9% decrease in DTP3 doses and a 10% decrease in measles doses delivered across the WHO AFRO region in April 2020(17), a smaller decrease than in other world regions. The reliance on outreach programmes during times of routine health service delivery, to obtain high immunisation coverage in many settings, may have mitigated the impact of the pandemic. The level of service disruption is also likely to be very context specific, and dependent on the local relationships between the health system and the community, local health system management and the budget available to conduct additional activities to mitigate impact. A study in Kinshasa found a reduction in measles vaccine coverage in 2 of 4 health zones, with larger hospitals more affected than health centres, but overall there was no difference in the number of vaccines delivered (18). We found no evidence of an impact on measles vaccine doses delivered across Goma. In Sierra Leone, data from two facilities in Bo region found a 32% decline in the number of ANC visits(19), whereas data from Kambia region showed no change in ANC. No impact on OPD visits for malaria was recorded at malaria reference centres in a variety of regions of Uganda(20), which contrasts with the 20-30% reduction observed in malaria OPD visits in Masaka. A strength of this analysis was that we focused resources on understanding service utilisation and provision in distinct settings and had the statistical power to detect effects if there were any.

There are several limitations of this analysis, the before-after design meant that we could not account for secular trends and there is evidence to suggest increasing catchment populations and increasing service activity across the three years in Goma. The lack of difference we observed between the pandemic periods compared to pre-pandemic periods may have been a reduction compared to what would be expected if 2020 had been ‘a normal year’. However, we did not have evidence of increased catchment populations or increasing activity levels over time for Masaka or Kambia. We assessed the comparability of the years of analysis with respect to climate factors and found no evidence of a difference in average maximum or minimum temperatures or atmospheric pressure between the years, by period. However, data were only available from international airport weather stations so do not account for local climate variations. We focused this analysis on government facilities so that recommendations were relevant to Ministry of Health officials; however, this means we do not have any evidence of whether the reductions in utilisation of government facilities coincided with an increase in the use of private or traditional health providers. Qualitative data in the DRC suggest the population may have opted to go to pharmacies, traditional practitioners, or private health structures in the belief that the state COVID testing and subsequent quarantine requirements, would be less strictly enforced.

We did not measure whether the vaccine doses were delivered via outreach or via routine services and therefore cannot estimate the extent to which service continuity relied on outreach during this time nor make any recommendations on continued outreach in these contexts, although health management stakeholders in Uganda suggest outreach mitigated some of the impact of the disruption to routine services. We assume catchment populations are relatively stable over time in order to compare utilisation across years with count data. There are limited data on population movement in and out of our study areas during the period of the study; however, some reports from Uganda indicate that there could have been population movement into Masaka from nearby cities in 2020, thus increasing the population, but this was transient and the effect was reported to last for only a few months (21, 22).

## Conclusion

We report evidence of disruptions to primary healthcare provision and utilization during the COVID-19 pandemic in three distinct settings: Goma, DRC; Kambia, Sierra Leone; and Masaka, Uganda. We found 20-50% fewer consultations for some essential primary healthcare services. The extent of the disruption and the services affected differed across these different settings. The level of disruption appeared to correlate with the strength of lockdown measures in the different settings and community attitudes towards the risk posed by COVID-19 especially in contexts with a history of Ebola outbreak responses. Mitigation strategies such as health communication campaigns and outreach services proved important to limit the impact of lockdowns on primary healthcare services.

## Data Availability

All data produced in the present study are available upon reasonable request to the authors

## Acknowledgements

We thank the data collection teams, the health facility staff and the Ministry of Health officials in the study countries for facilitating the collection of this data.

## Author contributions

**Conceived & designed the project**: K Gallagher, D Watson-Jones, B Greenwood, B Leigh, H Kavunga, E Ruzagira.

**Collected/ generated project data**: K Kasonia, D Tindanbil, J Kitonsa, F Zalwango, L Enria, A Mansaray, M James, Y Nije, D Tetsa Tata, B Lawal, A Drammeh, D Mukadi

**Contributed data/ analytic tools:** Kathy Baisley

**Performed the analysis**: K Gallagher

**Initial draft:** K. Kasonia, D Tindanbil

**Revise and review of all drafts:** K Kasonia, D Tindanbil, J Kitonsa, K Baisley, F Zalwango, L Enria, A Mansaray, M James, Y Nije, D Tetsa Tata, B Lawal, A Drammeh, B Lowe, D Mukadi, S Mounier-Jack, F Nakiyimba, P Obady, J Muhavi, B Greenwood, D Samai, B Leigh, D Watson-Jones, H Kavunga, E Ruzagira, K Gallagher

## References

1. World Health Organization. Pulse survey on continuity of essential health services during the COVID-19 pandemic. Interim report. 27 August. World Health Organization; 2020.

2. World Health Organization. Second round of national pulse survey on continuity of essential health services during the COVID-19 pandemic. 23rd April, 2021. 2021. Report No.: WHO/2019-nCoV/EHS_continuity/survey/2021.1.

3. Hale T, Angrist N, Goldszmidt R, Kira B, Petherick A, Phillips T, et al. A global panel database of pandemic policies (Oxford COVID-19 Government Response Tracker). Nature Human Behaviour. 2021;5(4):529–38.

4. Lim J, Broughan J, Crowley D, O’Kelly B, Fawsitt R, Burke MC, et al. COVID-19’s impact on primary care and related mitigation strategies: A scoping review. Eur J Gen Pract. 2021;27(1):166–75.

5. Vaccaro C, Mahmoud F, Aboulatta L, Aloud B, Eltonsy S. The impact of COVID-19 first wave national lockdowns on perinatal outcomes: a rapid review and meta-analysis. BMC Pregnancy and Childbirth. 2021;21(1):676.

6. McDonald HI, Tessier E, White JM, Woodruff M, Knowles C, Bates C, et al. Early impact of the coronavirus disease (COVID-19) pandemic and physical distancing measures on routine childhood vaccinations in England, January to April 2020. Euro surveillance : bulletin Europeen sur les maladies transmissibles = European communicable disease bulletin. 2020;25(19).

7. Miller HE, Henkel A, Leonard SA, Miller SE, Tran L, Bianco K, et al. The impact of the COVID-19 pandemic on postpartum contraception planning. American journal of obstetrics & gynecology MFM. 2021;3(5):100412–.

8. Wanyana D, Wong R, Hakizimana D. Rapid assessment on the utilization of maternal and child health services during COVID-19 in Rwanda. Public Health Action. 2021;11(1):12–21.

9. Bell D, Hansen KS, Kiragga AN, Kambugu A, Kissa J, Mbonye AK. Predicting the Impact of COVID-19 and the Potential Impact of the Public Health Response on Disease Burden in Uganda. Am J Trop Med Hyg. 2020;103(3):1191–7.

10. COVID-19; PfE-BRt. Effective implementation of Public Health and Social Measures in Uganda and DR Congo: Situational Analysis. Second data collection,19th August, 2020. Finding the Balance: Public Health and social measures in Uganda and DR Congo 3rd data collection. 2020.

11. The Government of Sierra Leone Ministry of Health & Sanitation. The Ministry of Health & Sanitation, Sierra Leone https://mohs.gov.sl/ [Last accessed 07April2022] 2022 [

12. The Government of Uganda. The Government of Uganda COVID-19 Response Information Hub https://covid19.gou.go.ug/timeline.html [last accessed 07April2022] 2022 [

13. Harris PA, Taylor R, Minor BL, Elliott V, Fernandez M, O’Neal L, et al. The REDCap consortium: Building an international community of software platform partners. J Biomed Inform. 2019;95:103208.

14. Harris PA, Taylor R, Thielke R, Payne J, Gonzalez N, Conde JG. Research electronic data capture (REDCap)--a metadata-driven methodology and workflow process for providing translational research informatics support. J Biomed Inform. 2009;42(2):377–81.

15. Hannah Ritchie, Edouard Mathieu, Lucas Rodés-Guirao, Cameron Appel, Charlie Giattino, Esteban Ortiz-Ospina, et al. “Coronavirus Pandemic (COVID-19)”. Published online at OurWorldInData.org. Retrieved from: ‘https://ourworldindata.org/coronavirus’ [Online Resource, last accessed 07April22] 2020 [

16. World Health Organization. Situation report: Ebola outbreak 2018-2020 - North Kivu-Ituri https://www.who.int/emergencies/situations/Ebola-2019-drc- [last accessed 07April2022] 2021 [

17. Shet A, Carr K, Danovaro-Holliday MC, Sodha SV, Prosperi C, Wunderlich J, et al. Impact of the SARS-CoV-2 pandemic on routine immunisation services: evidence of disruption and recovery from 170 countries and territories. The Lancet Global Health. 2022;10(2):e186–e94.

18. Hategeka C, Carter SE, Chenge FM, Katanga EN, Lurton G, Mayaka SM-N, et al. Impact of the COVID-19 pandemic and response on the utilisation of health services in public facilities during the first wave in Kinshasa, the Democratic Republic of the Congo. BMJ Global Health. 2021;6(7):e005955.

19. Aranda Z, Binde T, Tashman K, Tadikonda A, Mawindo B, Maweu D, et al. Disruptions in maternal health service use during the COVID-19 pandemic in 2020: experiences from 37 health facilities in low-income and middle-income countries. BMJ Global Health. 2022;7(1):e007247.

20. Namuganga JF, Briggs J, Roh ME, Okiring J, Kisambira Y, Sserwanga A, et al. Impact of COVID-19 on routine malaria indicators in rural Uganda: an interrupted time series analysis. Malaria journal. 2021;20(1):475.

21. Charles Onyango-Obbo. Urban-Rural migration opens up countryside to COVID-19 refugees. 09 May 2020 [https://www.theeastafrican.co.ke/tea/oped/comment/urban-rural-migration-opens-up-countryside-to-covid-19-refugees--1441052]. 2020.

22. UNHABITAT. COVID-19 through the Lens of Urban Rural Linkages - Guiding Principles and Framework for Action (URL-GP) [https://unhabitat.org/sites/default/files/2020/07/issue_brief_covid-19_through_the_lens_of_urban_rural_linkages_web_revised.pdf]. 2020.

