## Supplementary for "The impact of the COVID-19 pandemic on the provision & utilisation of primary health care services in Goma, Democratic Republic of the Congo, Kambia district, Sierra Leone & Masaka district, Uganda"

**Supplementary files**

**Goma, DRC**

Supplementary Figure 1. **Data Availability for each of the main outcomes, the number of facilities with data per week - Goma**

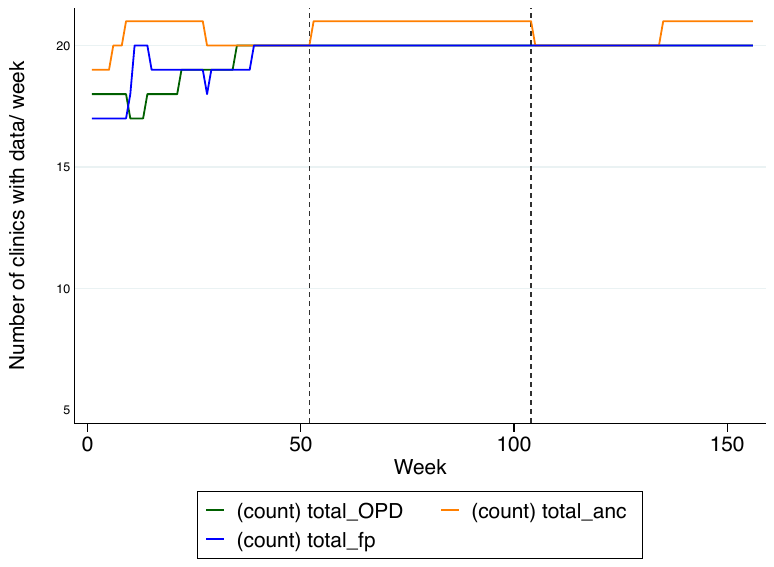

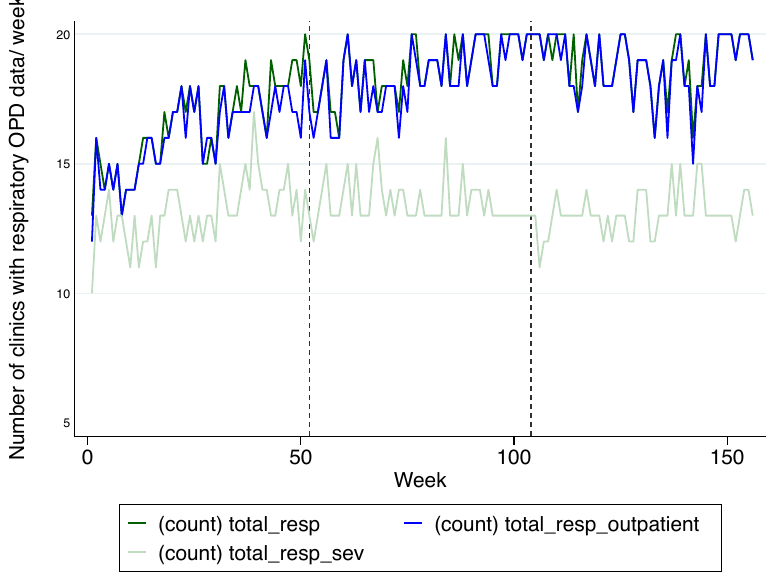

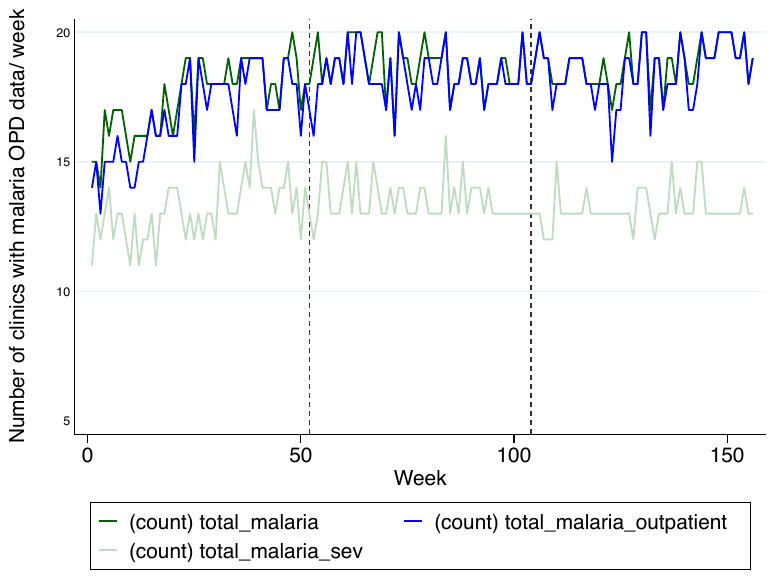

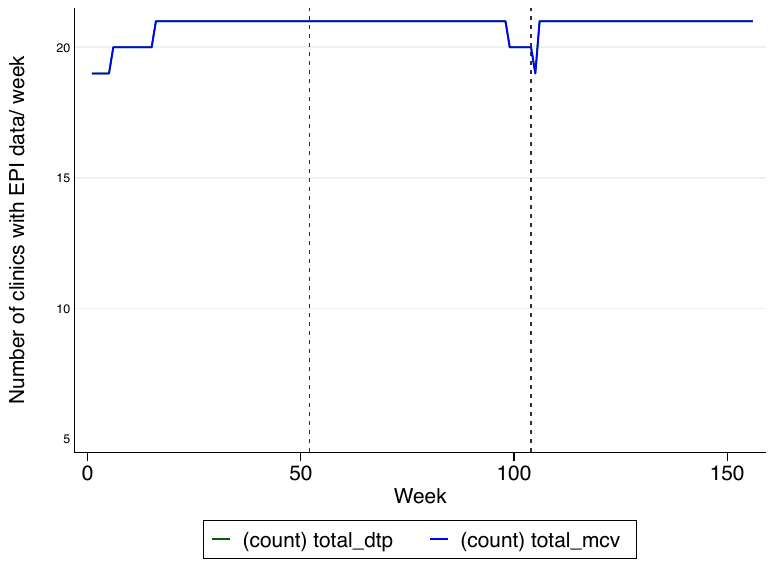

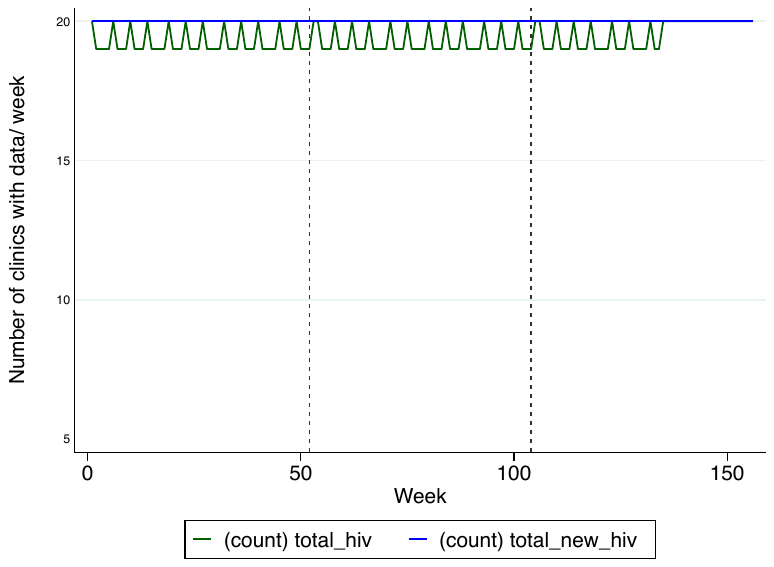

Abbreviations: ANC: Ante-natal care; ART: Anti-retroviral treatment; DTP: Diphtheria-tetanus-pertussis vaccine (3^rd^ dose)/ pentavalent vaccine (third dose); FP: Family Planning; HIV: Human Immunodeficiency Virus; MCV: Measles containing vaccine (1^st^ dose); OPD: under-5 outpatient department visit; Resp: respiratory outpatient department visits; TT: Tetanus toxoid vaccination

Supplementary Figure 2a. **Plots of the mean number of consultations per facility, per week for specific services, indicted on the y axis, across each calendar year of the study in 21 facilities in Goma.**

Vertical grey lines are the periods of analysis.

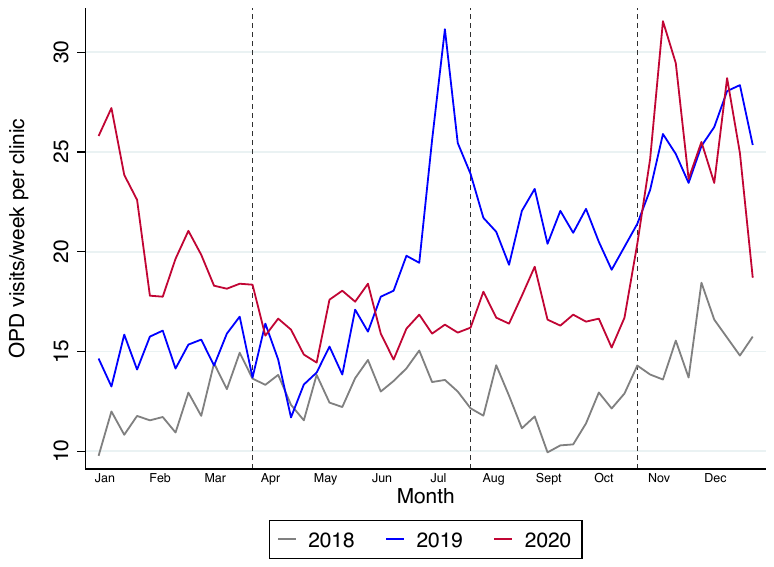

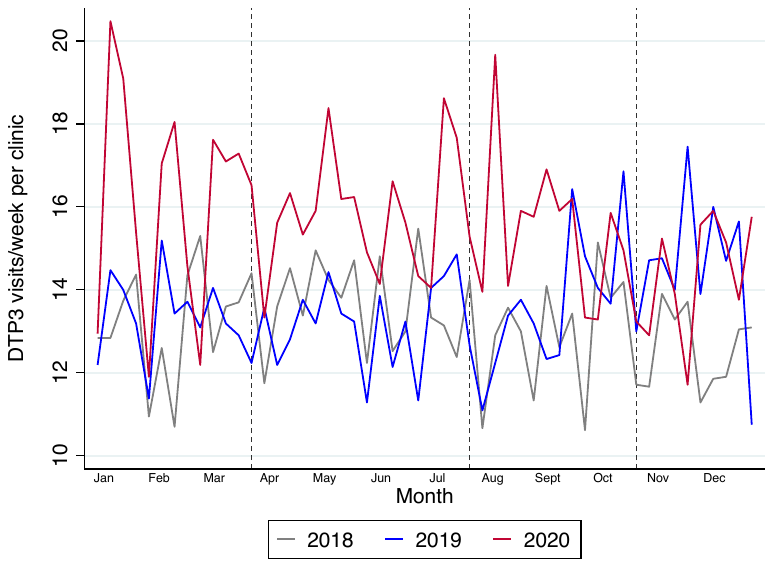

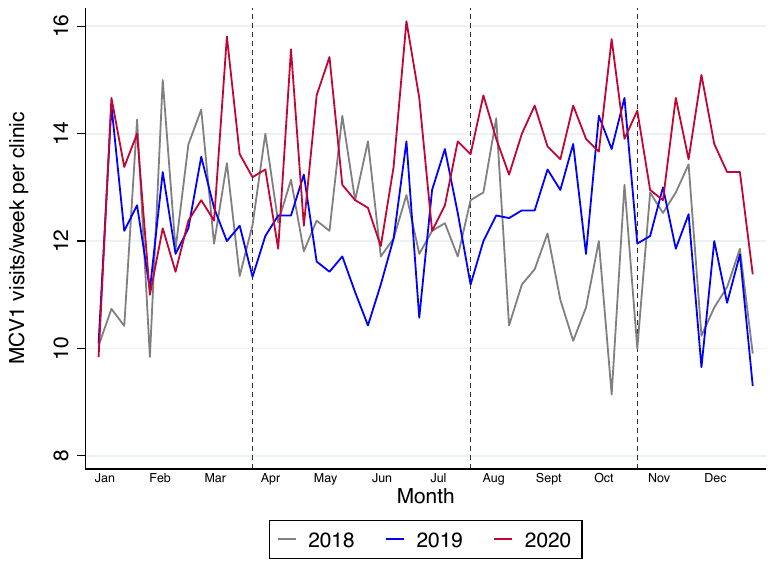

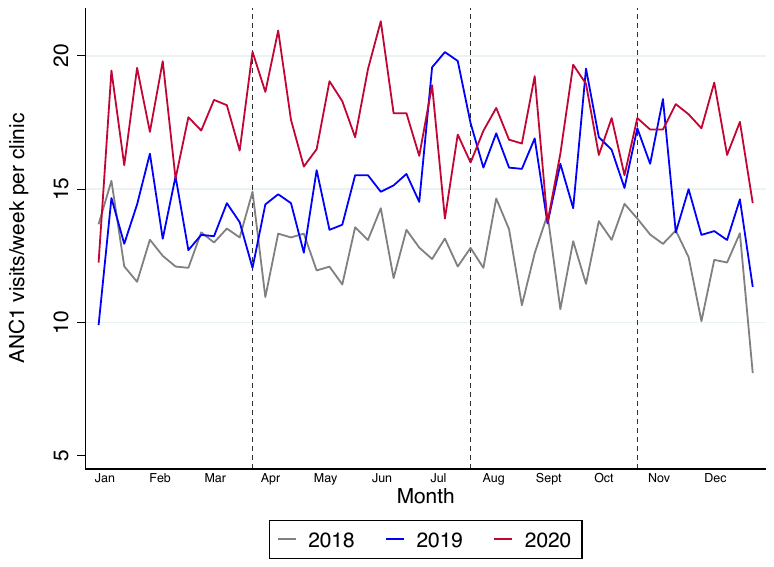

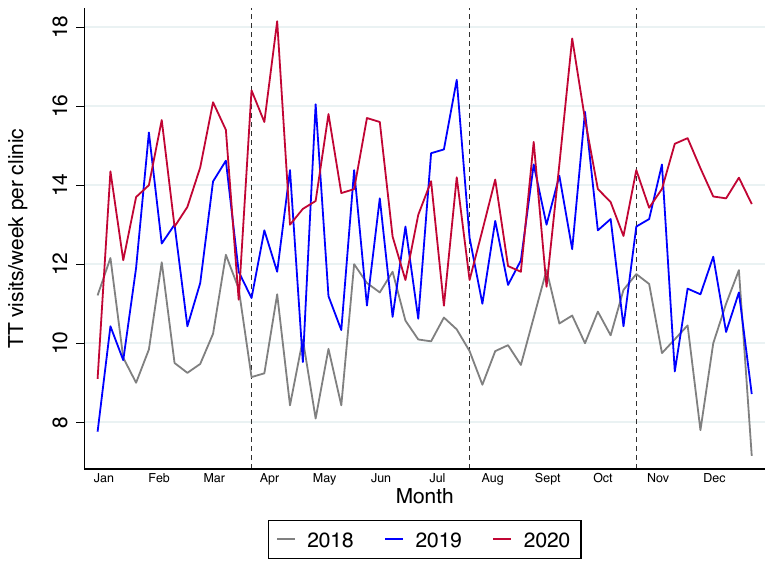

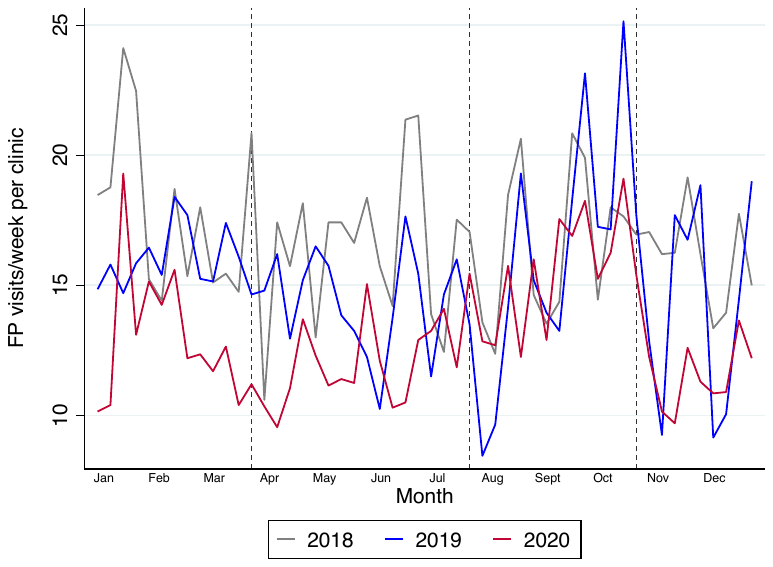

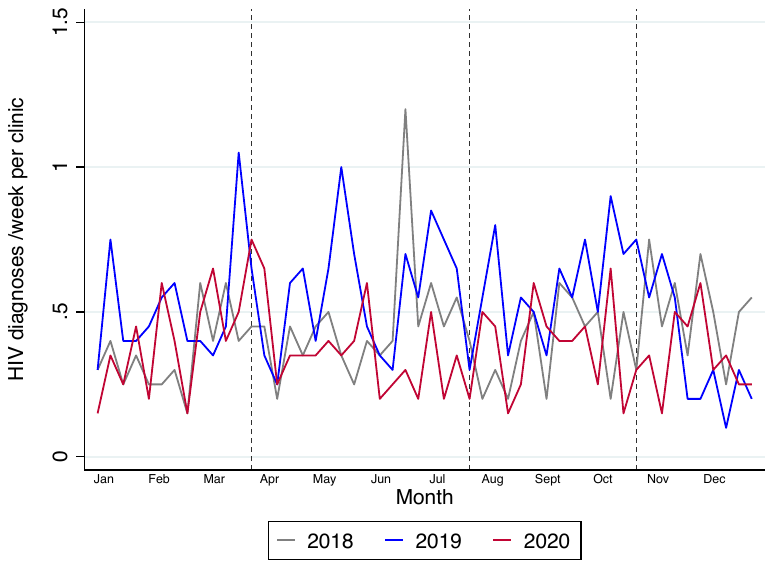

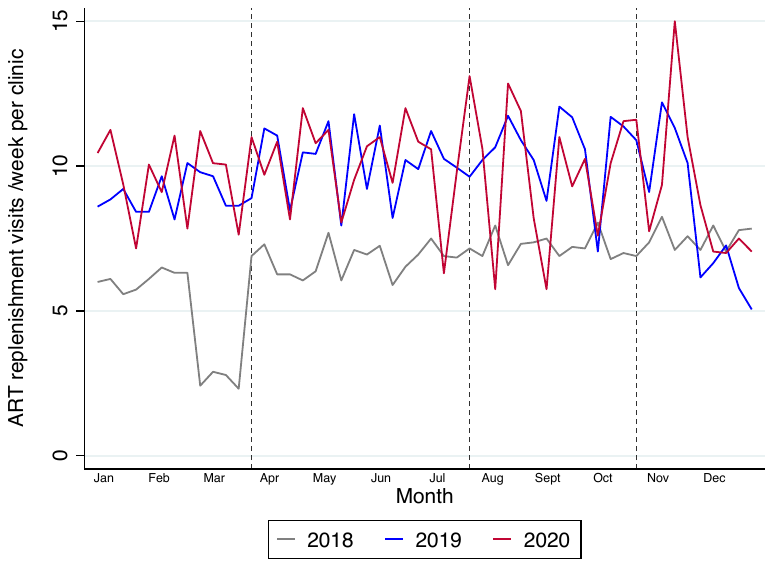

Abbreviations: ANC: Ante-natal care; ART: Anti-retroviral treatment; DTP: Diphtheria-tetanus-pertussis vaccine (3^rd^ dose)/ pentavalent vaccine (third dose); FP: Family Planning; HIV: Human Immunodeficiency Virus; MCV: Measles containing vaccine (1^st^ dose); OPD: under-5 outpatient department visit; Resp: respiratory outpatient department visits; TT: Tetanus toxoid vaccination

Supplementary Figure 2b. **Plots of the mean number of consultations per week for specific services, shown on the y axis combining an average of 2018-19 data and comparing this with 2020 data, in 21 facilities in Goma.**

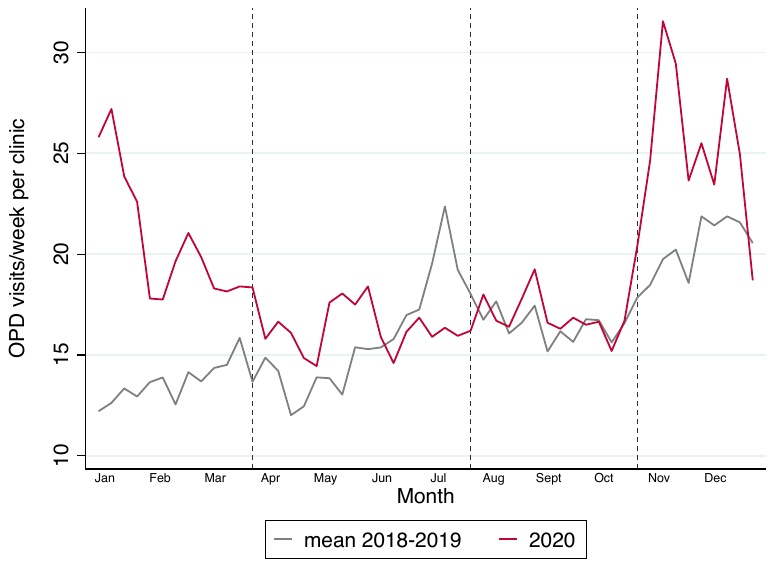

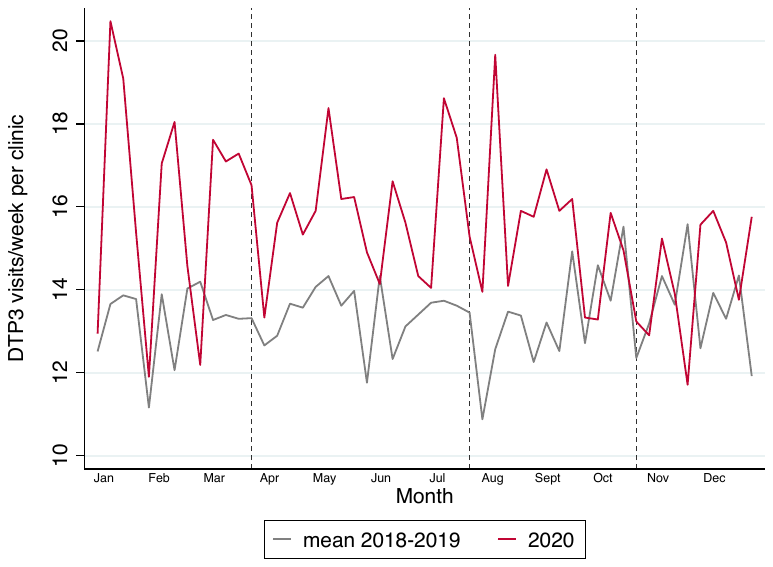

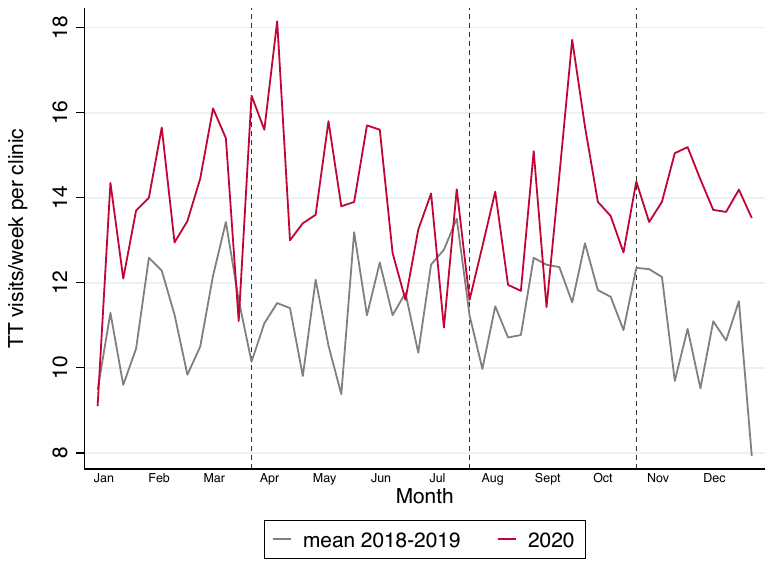

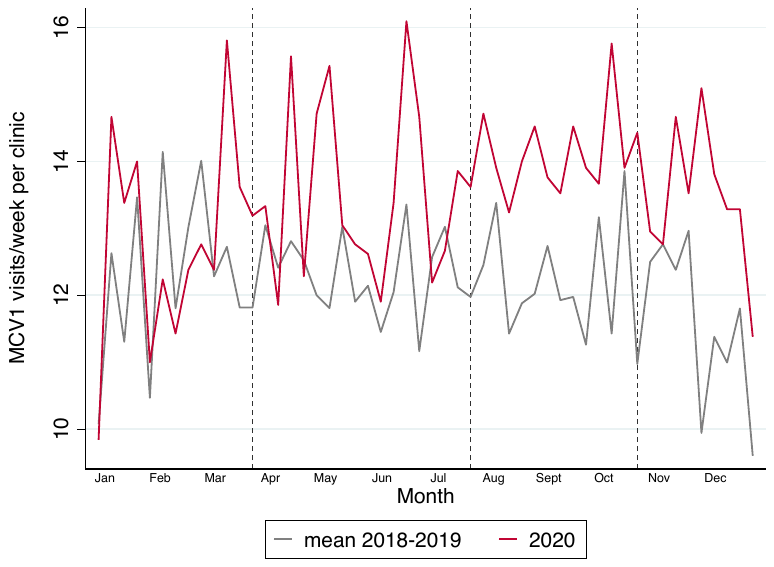

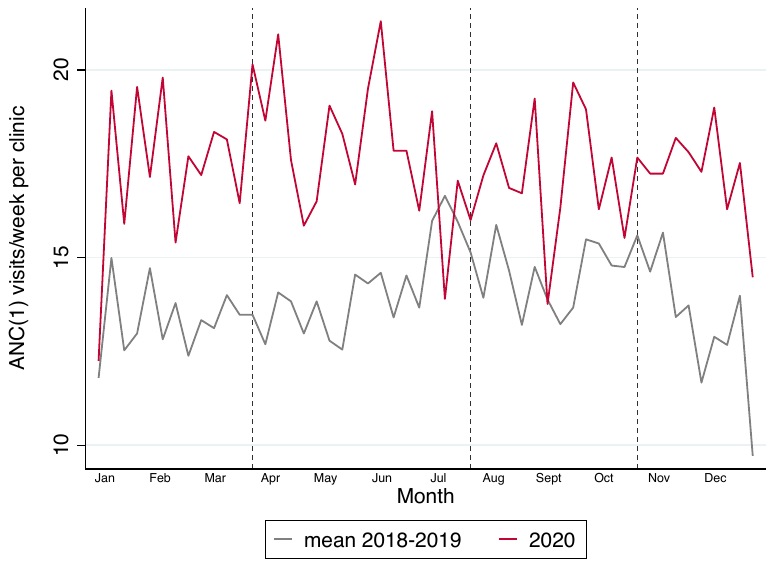

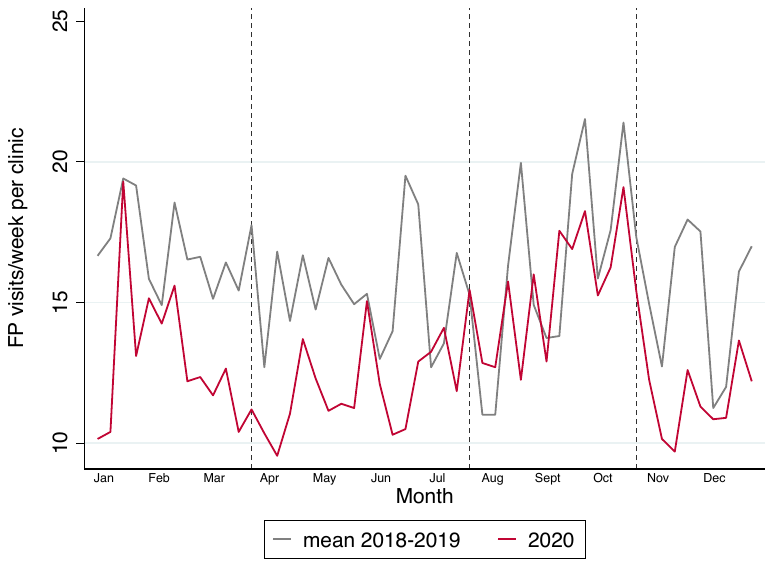

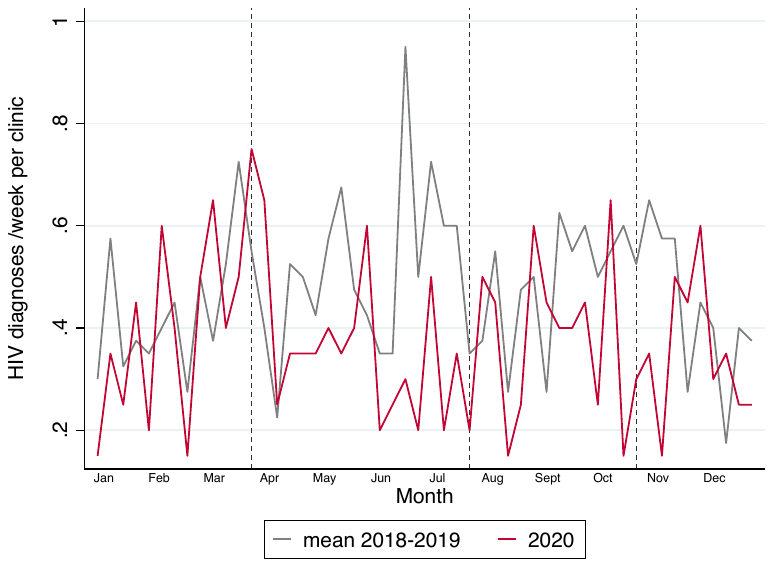

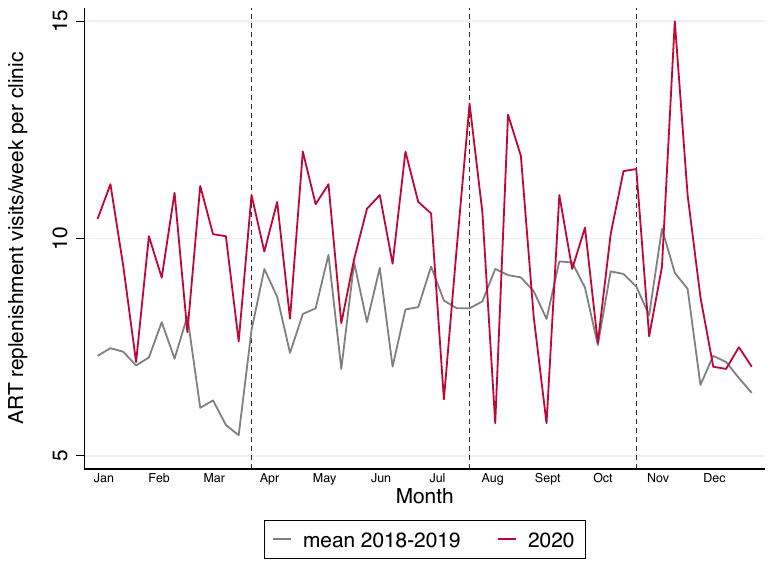

Supplementary Table 1: **Comparison of mean activity levels in similar calendar periods in 2018, 2019 and 2020 using a negative binomial regression model accounting for clustering by facility, and controlling for month (season) – in Goma**

|  |  | **01 January -22 March 2020**  **(pre-epidemic)** | | | **23 March – 19^th^ July 2020**  **Period 1 of lockdown** | | | | **20^th^ July – 18^th^ October 2020**  **Period 2 of lockdown** | | | | **19^th^ October – 27^th^ Dec 2020**  **Period 3 of lockdown** | | |
| --- | --- | --- | --- | --- | --- | --- | --- | --- | --- | --- | --- | --- | --- | --- | --- |
| **Outcome** | **Year** | **Mean visits/week (s.d)** | **ratio** | **p-value^2^** | **Mean visits/week (s.d)** | **ratio** | **p-value^2^** | **Mean visits/week (s.d)** | | **ratio** | **p-value^2^** | **Mean visits/week (s.d)** | | **ratio** | p-value^2^ |
| **OPD visits** | **2018** | 12.1 (13.0) | 0.80 (0.67-0.95) | <0.001 | 13.4 (11.7) | 0.76 (0.65-0.88) | 0.101 | 11.8 (11.2) | | 0.56 (0.43-0.73) | 0.004 | 15.2 (14.3) | | 0.61 (0.46-0.80) | 0.002 |
|  | **2019** | 15.1 (14.8) | 1 |  | 17.8 (17.1) | 1 |  | 21.3 (15.7) | | 1 |  | 25.2 (15.6) | | 1 |  |
|  | **2020** | 20.9 (17.6) | 1.38 (1.09-1.74) |  | 16.4 (17.4) | 0.94 (0.76-1.16) |  | 16.9 (17.7) | | 0.79 (0.60-1.04) |  | 25.1 (19.1) | | 1.00 (0.83-1.21) |  |
| **Resp OPD** | **2018** | 2.8 (2.7) | 0.74 (0.52-1.05) | 0.132 | 2.3 (2.3) | 0.66 (0.47-0.94) | 0.499 | 2.0 (2.6) | | 0.55 (0.39-0.78) | 0.544 | 3.8 (3.8) | | 0.55 (0.45-0.67) | <0.001 |
|  | **2019** | 3.8 (3.1) | 1 |  | 3.4 (3.3) | 1 |  | 3.5 (3.9) | | 1 |  | 7.1 (5.5) | | 1 |  |
|  | **2020** | 4.1 (3.9) | 1.04 (0.77-1.40) |  | 2.5 (2.9) | 0.74 (0.55-1.00) |  | 2.3 (3.2) | | 0.67 (0.43-1.05) |  | 8.0 (7.7) | | 1.15 (0.91-1.45) |  |
| **Severe resp** | **2018** | 0.96 (2.3) | 1.62 (0.91-2.87) | 0.608 | 0.7 (2.2) | 0.96 (0.38-2.43) | 0.231 | 0.5 (1.5) | | 0.93 (0.32-2.71) | 0.761 | 0.6 (1.4) | | 0.55 (0.32-0.98) | 0.068 |
|  | **2019** | 0.60 (1.10) | 1 |  | 0.7 (1.4) | 1 |  | 0.5 (1.1) | | 1 |  | 1.1 (1.7) | | 1 |  |
|  | **2020** | 0.70 (1.08) | 1.17 (0.57-2.40) |  | 0.3 (0.7) | 0.47 (0.28-0.78) |  | 0.4 (0.8) | | 0.75 (0.42-1.39) |  | 1.5 (2.2) | | 1.25 (0.78-2.00) |  |
| **Non-severe resp** | **2018** | 2.6 (2.6) | 0.73 (0.50-1.07) | 0.080 | 2.1 (2.4) | 0.72 (0.51-1.00) | 0.868 | 1.8 (2.2) | | 0.58 (0.40-0.85) | 0.537 | 3.7 (3.8) | | 0.61 (0.48-0.76) | <0.001 |
|  | **2019** | 3.5 (3.0) | 1 |  | 3.0 (3.0) | 1 |  | 3.0 (3.5) | | 1 |  | 6.3 (5.3) | | 1 |  |
|  | **2020** | 3.8 (4.2) | 1.07 (0.78-1.47) |  | 2.2 (2.4) | 0.74 (0.55-0.99) |  | 2.2 (3.1) | | 0.71 (0.44-1.14) |  | 7.0 (6.8) | | 1.14 (0.91-1.42) |  |
| **Malaria OPD** | **2018** | 4.9 (7.4) | 1.12 (0.77-1.63) | 0.460 | 4.6 (5.3) | 1.02 (0.85-1.22) | 0.767 | 3.6 (3.9) | | 0.64 (0.50-0.83) | 0.035 | 4.4 (4.6) | | 0.59 (0.39-0.89) | <0.001 |
|  | **2019** | 4.3 (5.4) | 1 |  | 4.5 (5.6) | 1 |  | 5.7 (5.9) | | 1 |  | 7.5 (7.1) | | 1 |  |
|  | **2020** | 5.9 (6.6) | 1.36 (0.91-2.02) |  | 4.8 (6.3) | 1.06 (0.83-1.36) |  | 5.1 (6.1) | | 0.89 (0.76-1.05) |  | 7.5 (7.2) | | 1.01 (0.81-1.25) |  |
| **Severe malaria** | **2018** | 5.7 (16.6) | 1.46 (1.06-2.02) | 0.230 | 4.0 (11.5) | 0.94 (0.78-1.14) | <0.001 | 2.5 (7.1) | | 0.54 (0.43-0.67) | <0.001 | 2.3 (6.3) | | 0.46 (0.36-0.60) | <0.001 |
|  | **2019** | 3.9 (10.4) | 1 |  | 4.2 (11.5) | 1 |  | 4.7 (12.6) | | 1 |  | 4.9 (12.3) | | 1 |  |
|  | **2020** | 4.6 (11.9) | 1.19 (1.04-1.36) |  | 5.4 (14.6) | 1.28 (1.17-1.41) |  | 5.4 (13.8) | | 1.14 (1.01-1.29) |  | 4.7 (11.5) | | 0.97 (0.79-1.19) |  |
| **Non-severe malaria** | **2018** | 4.4 (7.4) | 1.17 (0.73-1.87) | 0.701 | 4.3 (6.9) | 1.09 (0.85-1.41) | 0.792 | 3.1 (3.5) | | 0.64 (0.46-0.89) | 0.081 | 4.0 (4.3) | | 0.61 (0.39-0.96) | 0.005 |
|  | **2019** | 3.8 (4.9) | 1 |  | 3.9 (5.2) | 1 |  | 4.8 (5.4) | | 1 |  | 6.6. (6.7) | | 1 |  |
|  | **2020** | 5.0 (6.2) | 1.31 (0.84-2.06) |  | 4.1 (6.1) | 1.04 (0.77-1.39) |  | 4.4 (5.9) | | 0.91 (0.75-1.11) |  | 6.5 (6.8) | | 0.99 (0.82-1.19) |  |
| **DTP** | **2018** | 13.1 (12.0) | 0.98 (0.86-1.11) | 0.015 | 13.7 (11.1) | 1.04 (0.92-1.18) | 0.076 | 13.0 (9.3) | | 0.96 (0.85-1.08) | 0.024 | 12.5 (9.5) | | 0.87 (0.74-1.02) | 0.109 |
|  | **2019** | 13.4 (10.6) | 1 |  | 13.2 (10.1) | 1 |  | 13.6 (10.5) | | 1 |  | 14.5 (10.3) | | 1 |  |
|  | **2020** | 16.1 (12.7) | 1.21 (1.07-1.36) |  | 15.9 (12.0) | 1.20 (1.07-1.36) |  | 15.5 (10.8) | | 1.14 (0.98-1.32) |  | 14.3 (10.0) | | 0.99 (0.83-1.18) |  |
| **MCV** | **2018** | 12.3 (10.2) | 0.99 (0.86-1.14) | 0.643 | 12.6 (10.0) | 1.04 (0.90-1.21) | 0.406 | 11.6 (9.3) | | 0.90 (0.80-1.01) | 0.021 | 11.6 (8.7) | | 1.01 (0.90-1.13) | 0.038 |
|  | **2019** | 12.4 (9.0) | 1 |  | 12.0 (8.4) | 1 |  | 12.9 (7.9) | | 1 |  | 11.5 (7.5) | | 1 |  |
|  | **2020** | 12.8 (8.6) | 1.04 (0.87-1.23) |  | 13.5 (8.8) | 1.12 (0.98-1.28) |  | 14.1 (8.6) | | 1.09 (0.98-1.21) |  | 13.5 (8.3) | | 1.18 (1.03-1.34) |  |
| **ANC** | **2018** | 13.0 (11.6) | 0.95 (0.85-1.06) | 0.005 | 12.8 (11.9) | 0.83 (0.77-0.91) | <0.001 | 12.8 (11.0) | | 0.79 (0.65-0.96) | 0.001 | 12.2 (11.1) | | 0.84 (0.68-1.04) | 0.004 |
|  | **2019** | 13.7 (12.0) | 1 |  | 15.4 (13.4) | 1 |  | 16.2 (13.8) | | 1 |  | 14.6 (13.1) | | 1 |  |
|  | **2020** | 17.3 (14.2) | 1.26 (1.07-1.49) |  | 18.0 (14.7) | 1.17 (1.04-1.32) |  | 17.1 (14.9) | | 1.05 (0.90-1.23) |  | 17.3 (14.4) | | 1.19 (1.02-1.38) |  |
| **TT** | **2018** | 10.5 (9.7) | 0.88 (0.76-1.03) | 0.006 | 10.1 (9.1) | 0.80 (0.73-0.87) | <0.001 | 10.3 (8.5) | | 0.80 (0.67-0.97) | <0.022 | 10.1 (8.2) | | 0.88 (0.72-1.09) | 0.002 |
|  | **2019** | 11.9 (10.8) | 1 |  | 12.8 (11.9) | 1 |  | 12.8 (10.2) | | 1 |  | 11.5 (10.2) | | 1 |  |
|  | **2020** | 13.5 (11.1) | 1.14 (0.95-1.36) |  | 14.2 (11.8) | 1.12 (0.95-1.31) |  | 13.6 (12.7) | | 1.06 (0.88-1.28) |  | 14.1 (12.1) | | 1.23 (1.00-1.53) |  |
| **FP** | **2018** | 17.5 (16.7) | 1.08 (0.85-1.38) | 0.084 | 16.6 (18.2) | 1.15 (0.99-1.34) | 0.005 | 16.6 (17.8) | | 1.05 (0.79-1.40) | 0.637 | 16.2 (17.2) | | 1.11 (0.81-1.52) | 0.055 |
|  | **2019** | 16.1 (16.0) | 1 |  | 14.4 (17.0) | 1 |  | 16.0 (25.9) | | 1 |  | 14.6 (23.8) | | 1 |  |
|  | **2020** | 13.1 (16.8) | 0.81 (0.65-1.01) |  | 11.9 (11.8) | 0.83 (0.69-0.98) |  | 15.5 (15.9) | | 0.98 (0.69-1.37) |  | 11.9 (11.2) | | 0.82 (0.56-1.18) |  |
| **ART visits** | **2018** | 4.9 (15.1) | 0.54 (0.37-0.78) | 0.006 | 6.8 (18.3) | 0.67 (0.40-1.10) | 0.213 | 7.2 (18.7) | | 0.69 (0.42-1.12) | 0.287 | 7.5 (18.9) | | 0.90 (0.39-2.09) | 0.594 |
|  | **2019** | 9.0 (18.9) | 1 |  | 10.1 (20.6) | 1 |  | 10.5 (22.1) | | 1 |  | 8.5 (19.0) | | 1 |  |
|  | **2020** | 9.6 (21.7) | 1.06 (0.73-1.55) |  | 10.1 (21.8) | 1.00 (0.78-1.27) |  | 9.8 (22.9) | | 0.94 (0.79-1.11) |  | 9.2 (19.1) | | 1.09 (0.84-1.42) |  |
| **New HIV** | **2018** | 0.35 (0.81) | 0.69 (0.40-1.20) | 0.782 | 0.46 (1.35) | 0.80 (0.46-1.38) | 0.526 | 0.38 (1.16) | | 0.67 (0.34-1.35) | 0.940 | 0.50 (1.26) | | 1.31 (0.62-2.75) | 0.256 |
|  | **2019** | 0.51 (1.18) | 1 |  | 0.58 (1.33) | 1 |  | 0.57 (1.45) | | 1 |  | 0.39 (1.31) | | 1 |  |
|  | **2020** | 0.38 (0.83) | 0.75 (0.53-1.07) |  | 0.38 (0.97) | 0.66 (0.53-0.80) |  | 0.38 (0.99) | | 0.66 (0.44-0.99) |  | 0.35 (0.91) | | 0.92 (0.52-1.65) |  |

^2^ The document p-values are Wald tests, testing the hypothesis that the coefficients for ‘year’ estimated by the negative binomial regression model, are equal. Likelihood ratio tests are invalid given that the likelihood estimated under robust standard errors does not account for clustering: <https://www.stata.com/support/faqs/statistics/likelihood-ratio-test/> . Abbreviations: ANC: Ante-natal care; ART: Anti-retroviral treatment; DTP: Diphtheria-tetanus-pertussis vaccine (3^rd^ dose)/ pentavalent vaccine (third dose); FP: Family Planning; HIV: Human Immunodeficiency Virus; MCV: Measles containing vaccine (1^st^ dose); OPD: under-5 outpatient department visit; Resp: respiratory outpatient department visits; TT: Tetanus toxoid vaccination

**Kambia, Sierra Leone**

Supplementary Figure 3. **Data Availability for each of the main outcomes, the number of facilities with data per week – Kambia, Sierra Leone**

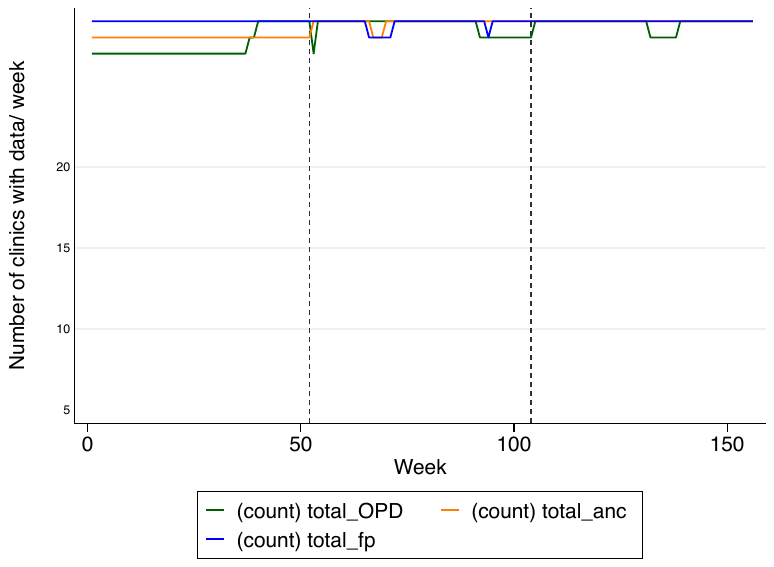

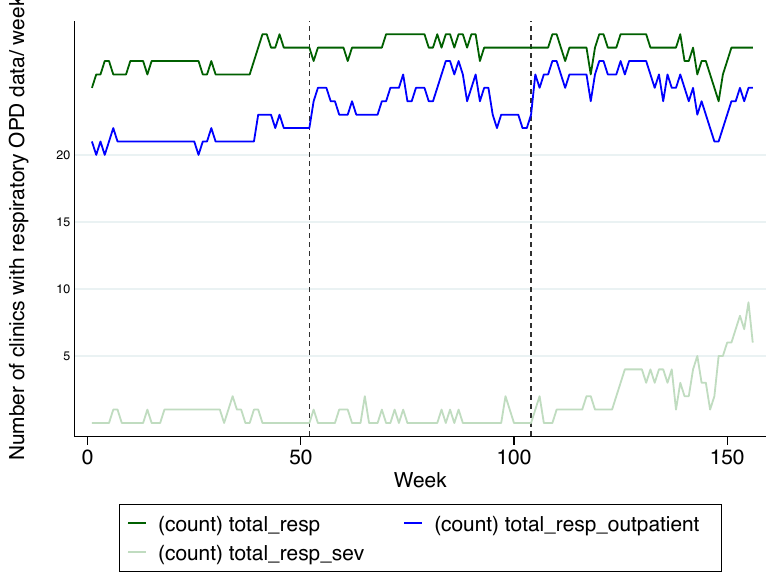

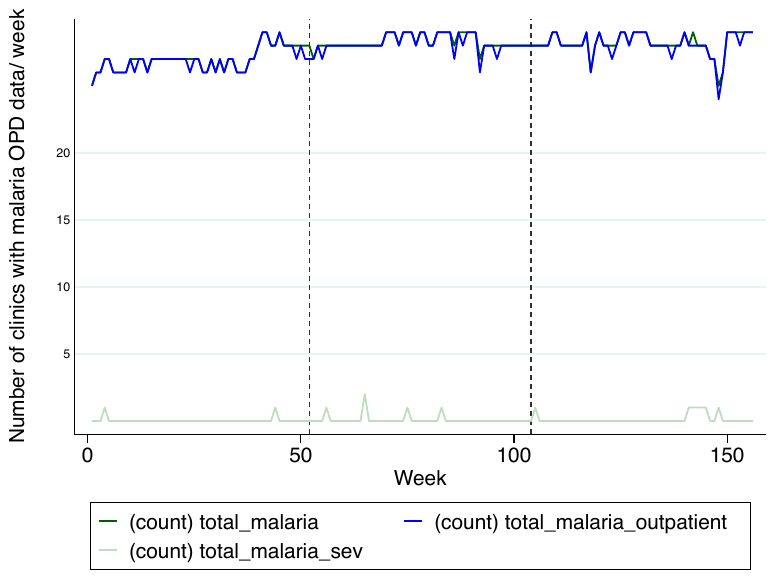

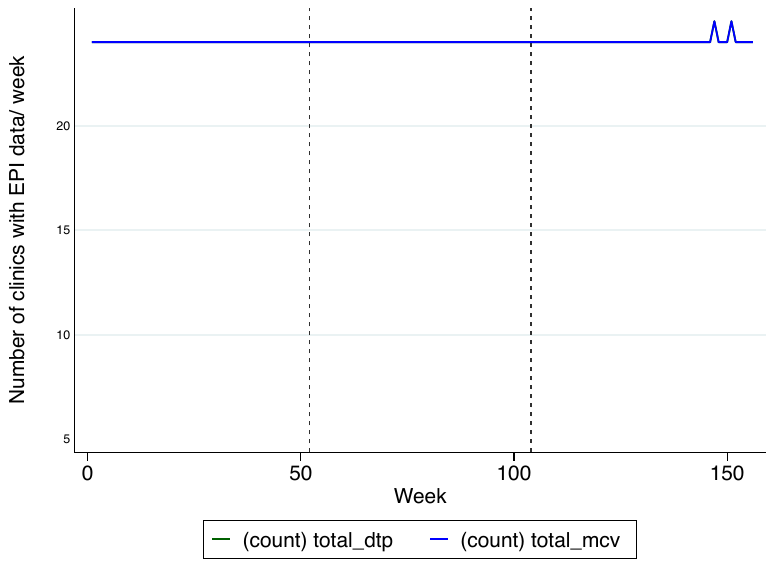

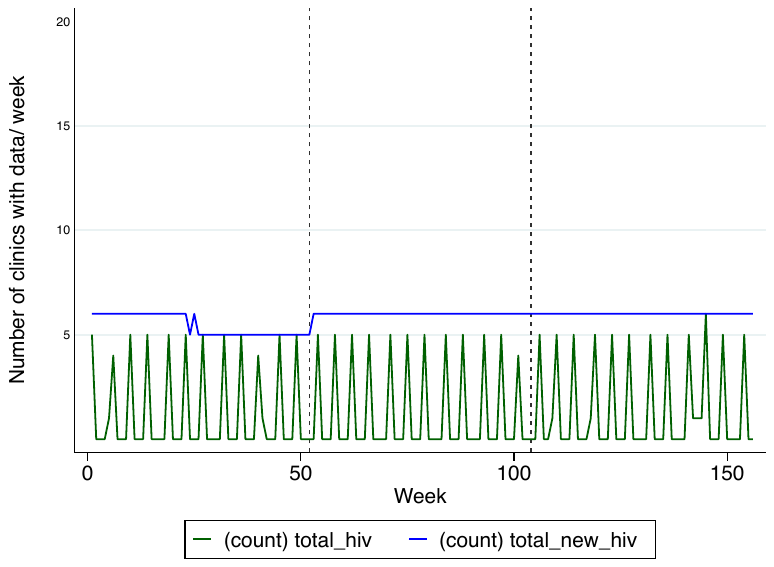

*missing data for severity, and outpatient data for resp/malaria

Abbreviations: ANC: Ante-natal care; ART: Anti-retroviral treatment; DTP: Diphtheria-tetanus-pertussis vaccine (3^rd^ dose)/ pentavalent vaccine (third dose); FP: Family Planning; HIV: Human Immunodeficiency Virus; MCV: Measles containing vaccine (1^st^ dose); OPD: under-5 outpatient department visit; Resp: respiratory outpatient department visits; TT: Tetanus toxoid vaccination

Supplementary Figure 4a. **Plots of the mean number of consultations per facility, per week for specific services, indicted on the y axis, across each calendar year of the study in 29 facilities in Kambia district, Sierra Leone.** Vertical grey lines are the periods of analysis.

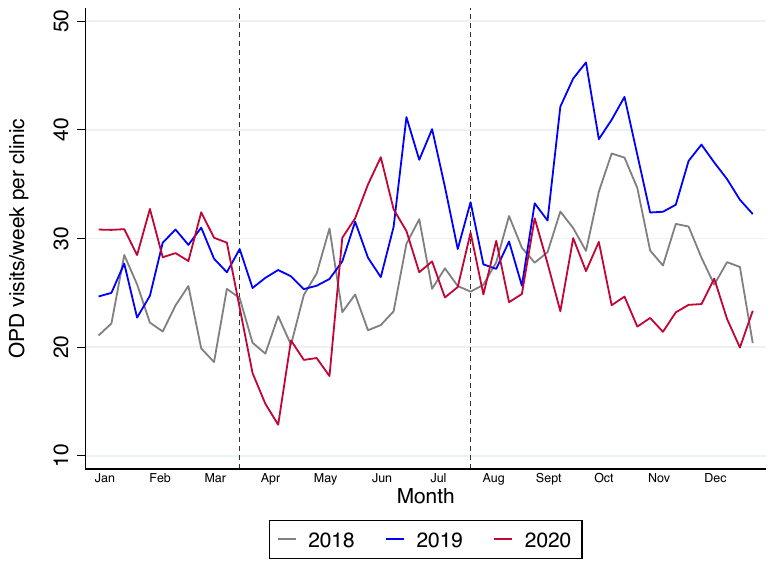

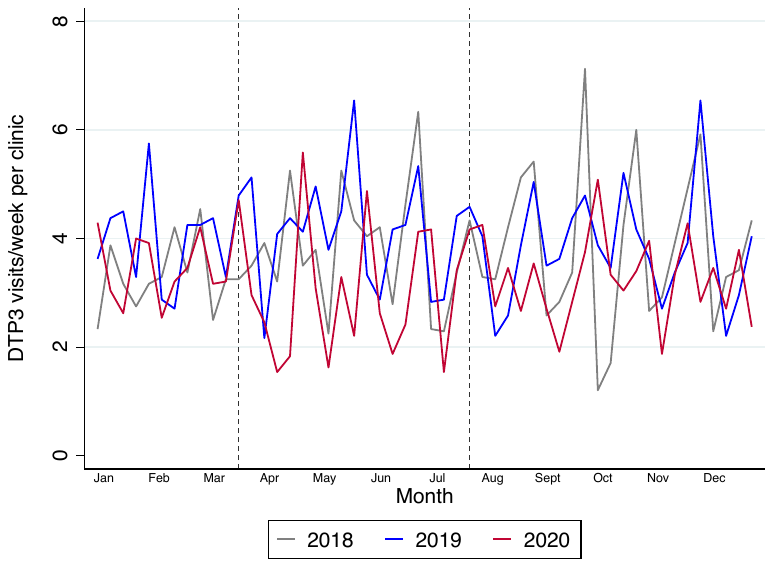

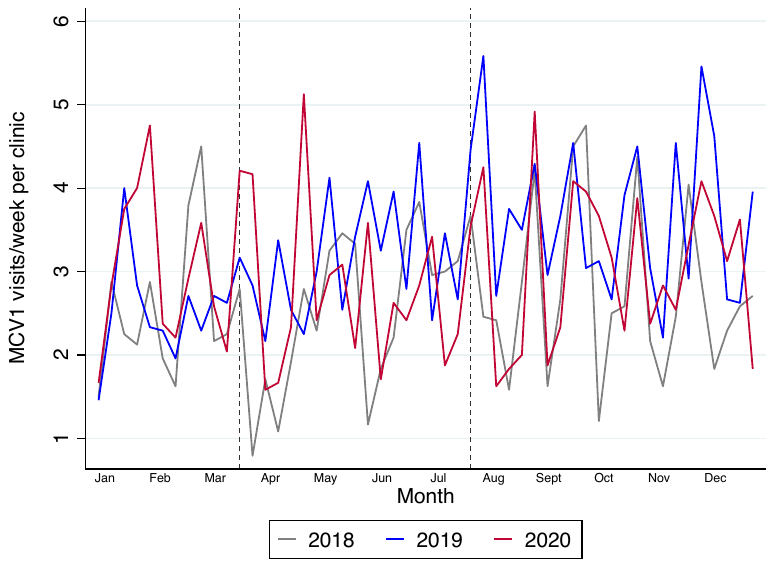

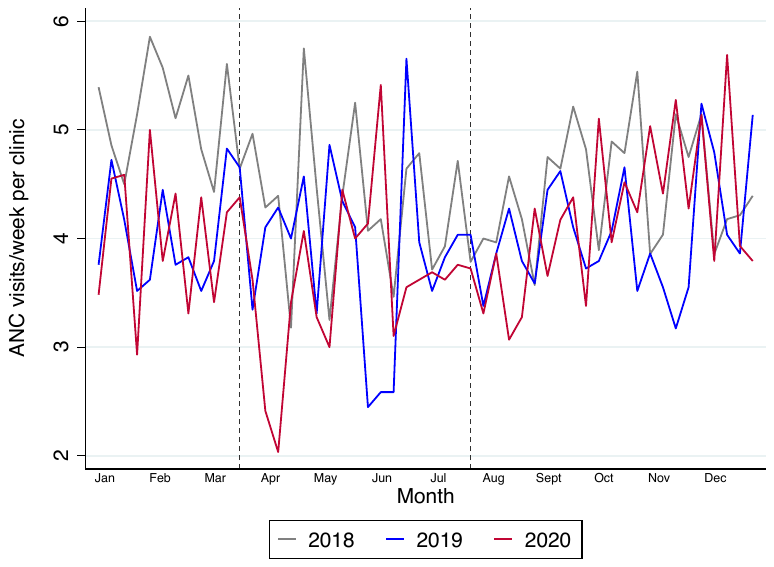

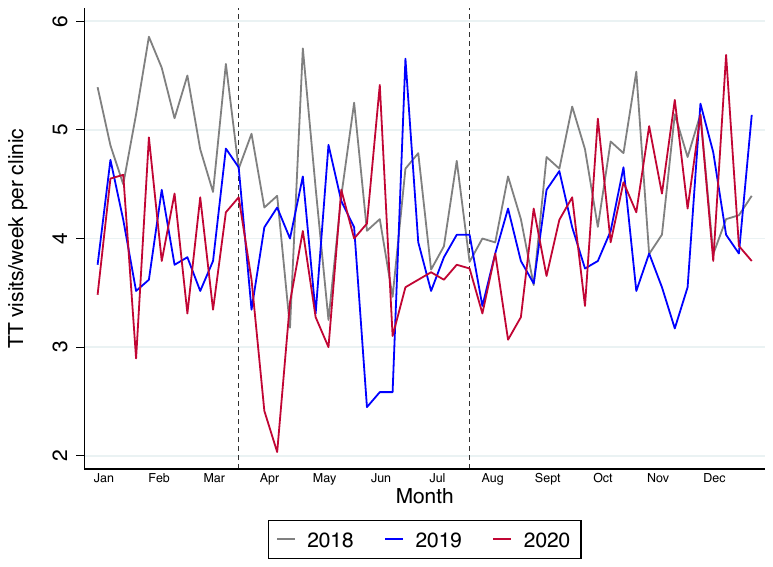

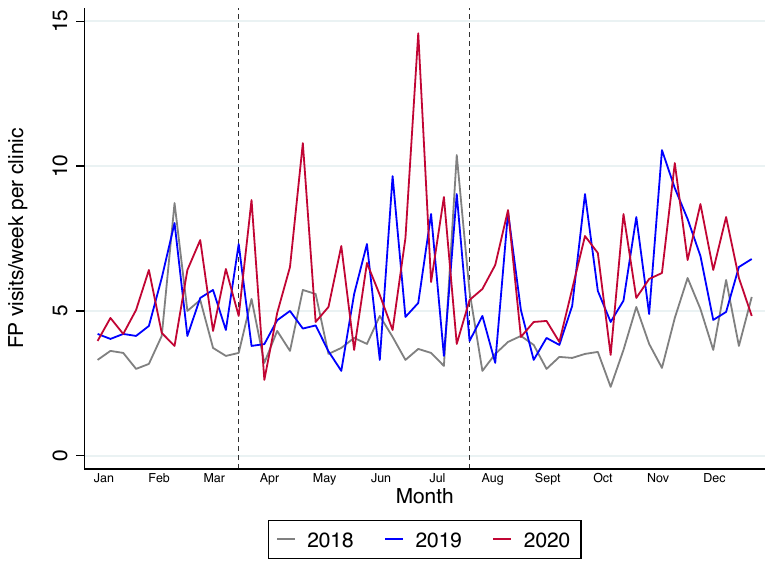

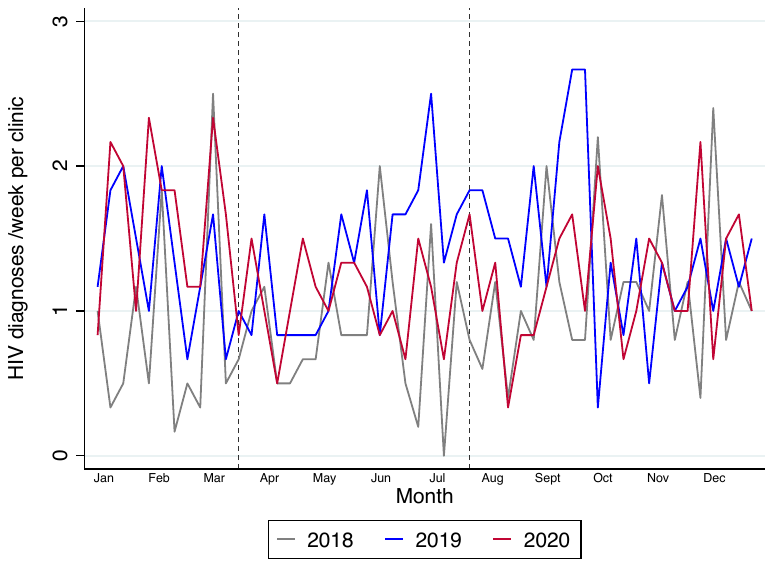

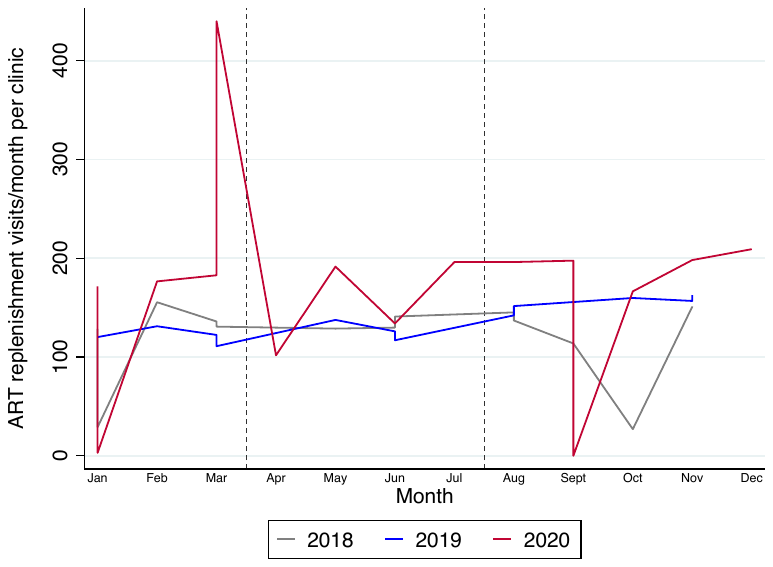

Supplementary Figure 4b. **Mean number of consultations per facility per week, combining an average of 2018-19 data and comparing this with 2020 data, in 29 facilities in Kambia district, Sierra Leone.**

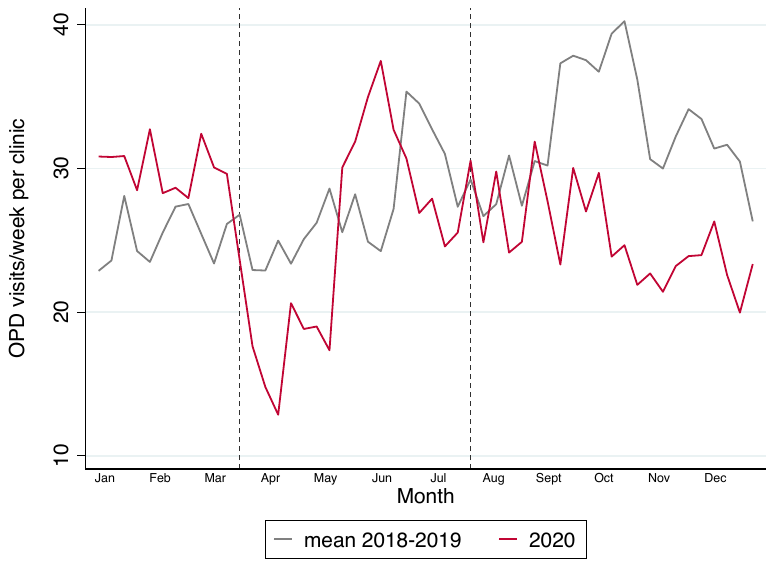

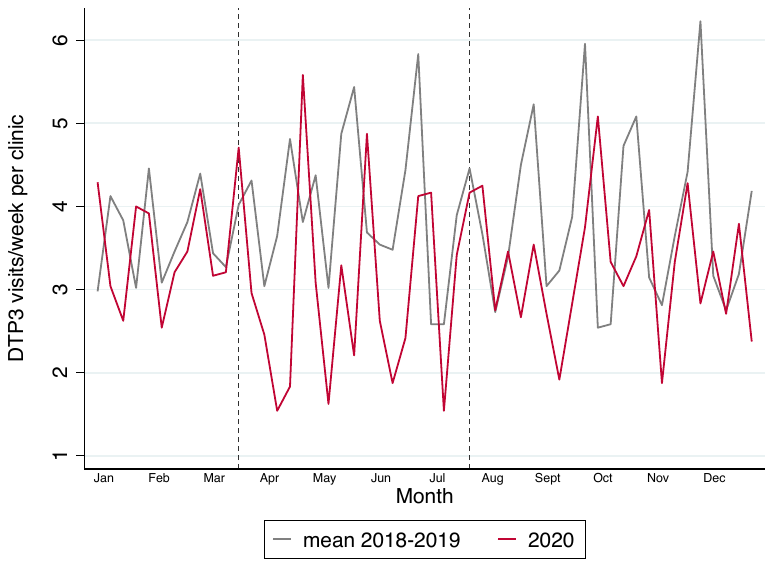

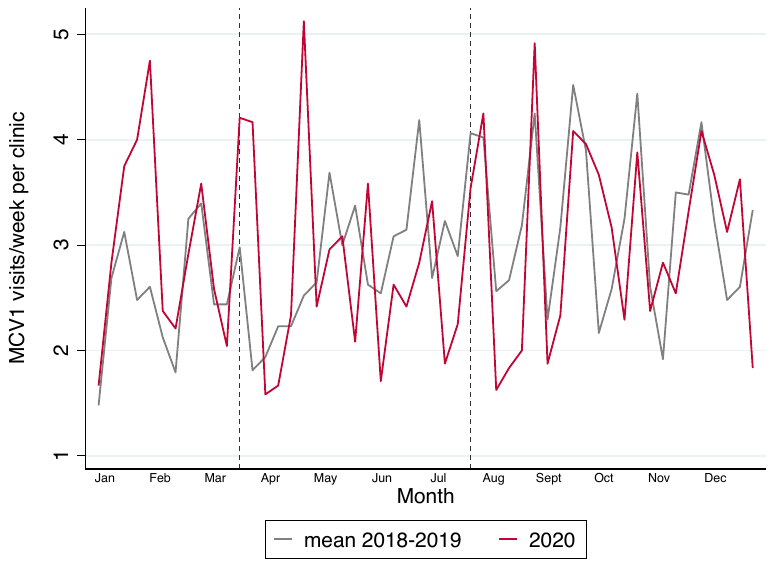

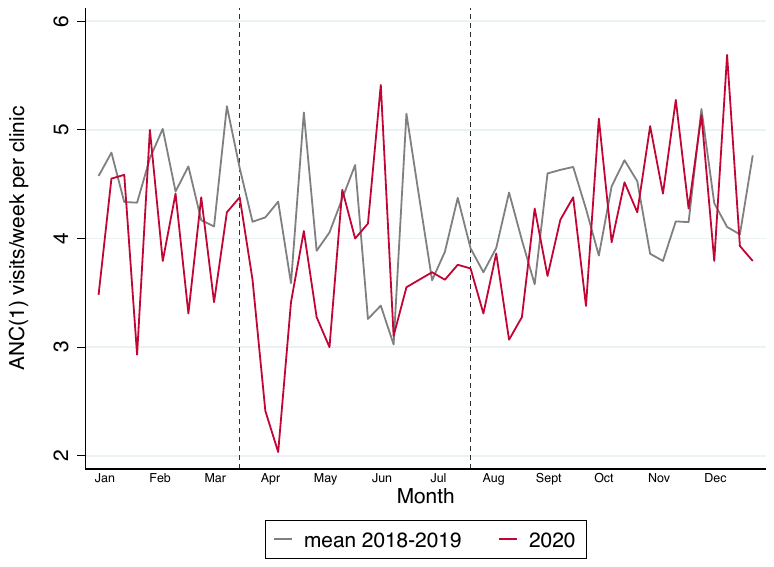

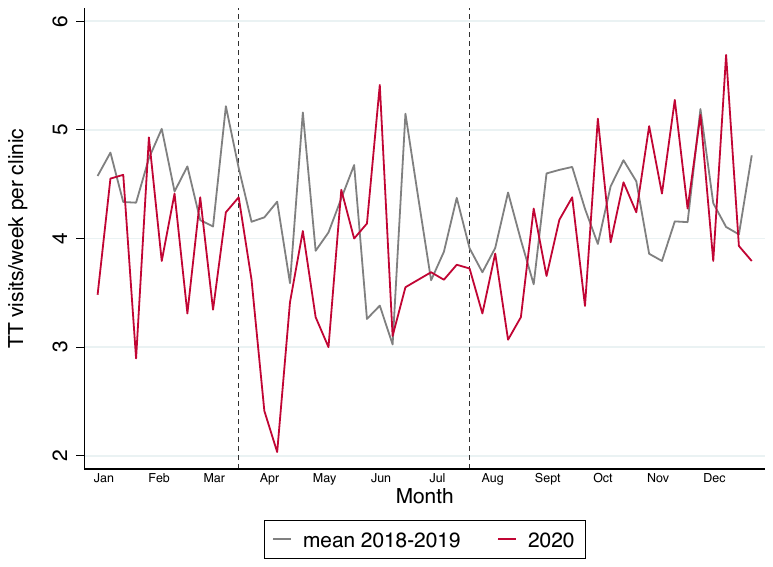

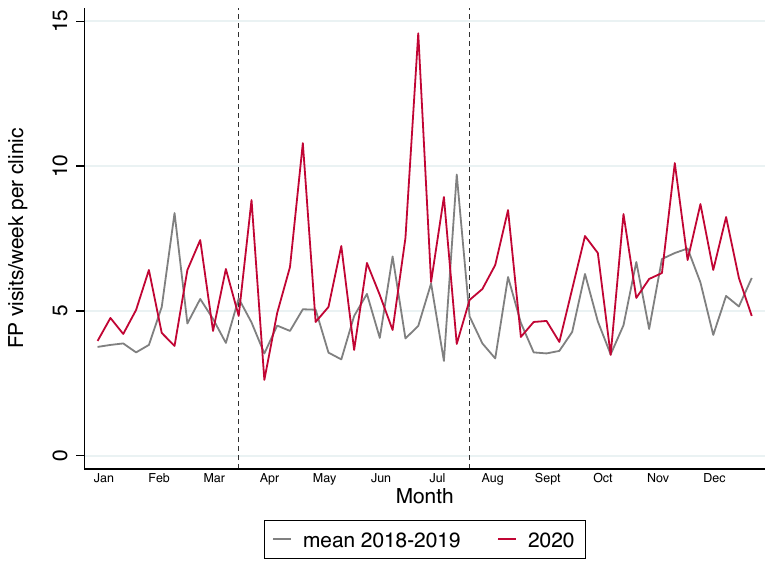

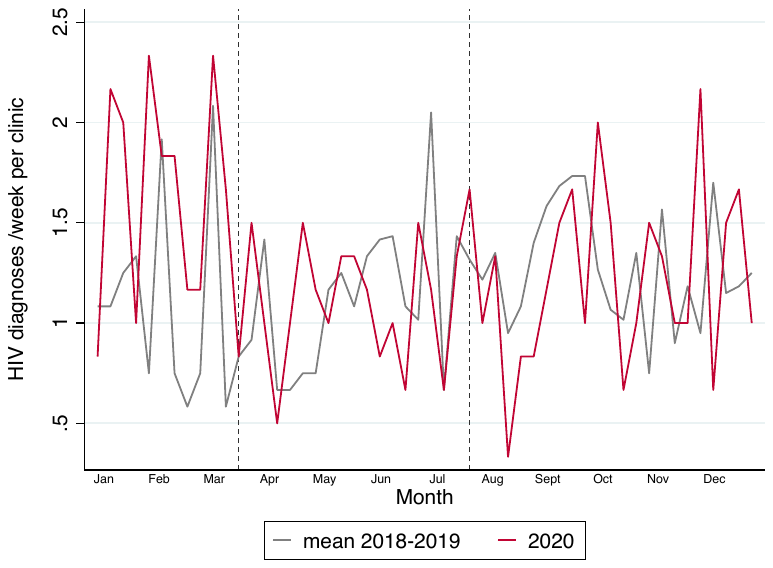

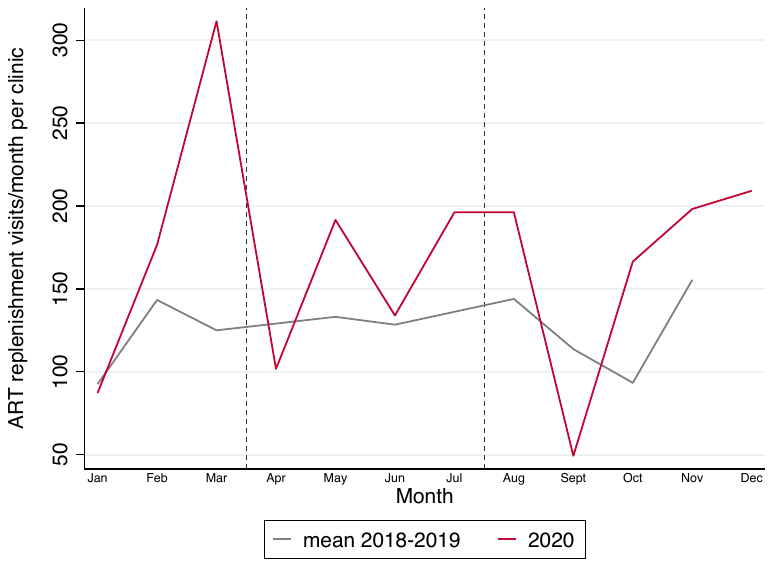

Supplementary Table 2: **Comparison of mean activity levels in similar calendar periods in 2018, 2019 and 2020 using a negative binomial regression model accounting for clustering by facility, and controlling for month (season) – in Kambia, Sierra Leone**

|  |  | **01Jan-22Mar**  **(pre-pandemic)** | | | **23 March 2020 – 19^th^ July 2020**  **Period 1 of lockdown** | | | **20^th^ July 2020 – 27^th^ Dec 2020**  **Period 2 of lockdown** | | |
| --- | --- | --- | --- | --- | --- | --- | --- | --- | --- | --- |
| **Outcome** | **Year** | **Mean visits/week (s.d)** | **Mean ratio** | **p-value^2^** | **Mean visits/week (s.d)** | **Mean ratio** | **p-value^2^** | **Mean visits/week (s.d)** | **Mean ratio** | **p-value^2^** |
| **OPD visits** | **2018** | 23.1 (11.8) | 0.85 (0.70-1.03) | 0.014 | 24.7 (13.5) | 0.83 (0.72-0.94) | 0.946 | 29.7 (16.3) | 0.84 (0.69-1.01) | 0.066 |
|  | **2019** | 27.4 (16.3) | 1 |  | 30.0 (17.1) | 1 |  | 35.4 (18.9) | 1 |  |
|  | **2020** | 30.1 (19.6) | 1.10 (0.92-1.31) |  | 24.9 (16.8) | 0.82 (0.72-0.94) |  | 25.3 (14.4) | 0.71 (0.64-0.80) |  |
| **Resp OPD** | **2018** | 12.4 (7.9) | 0.72 (0.55-0.95) | 0.214 | 13.4 (9.9) | 0.79 (0.66-0.95) | 0.021 | 18.7 (12.6) | 1.09 (0.89-1.33) | <0.001 |
|  | **2019** | 17.2 (14.3) | 1 |  | 16.9 (11.4) | 1 |  | 17.2 (10.7) | 1 |  |
|  | **2020** | 14.4 (9.6) | 0.84 (0.67-1.06) |  | 10.2 (9.2) | 0.60 (0.49-0.74) |  | 11.4 (10.2) | 0.66 (0.55-0.80) |  |
| **Malaria OPD** | **2018** | 12.8 (8.1) | 0.75 (0.62-0.90) | 0.002 | 15.7 (10.9) | 0.81 (0.70-0.94) | 0.852 | 20.1 (13.5) | 0.94 (0.80-1.10) | 0.021 |
|  | **2019** | 17.2 (11.1) | 1 |  | 19.5 (13.2) | 1 |  | 21.4 (12.6) | 1 |  |
|  | **2020** | 18.0 (12.1) | 1.05 (0.86-1.28) |  | 16.2 (11.7) | 0.83 (0.72-0.96) |  | 16.2 (9.5) | 0.76 (0.68-0.85) |  |
| **DTP** | **2018** | 3.3 (4.7) | 0.84 (0.59-1.21) | 0.852 | 3.8 (5.8) | 0.92 (0.73-1.14) | 0.189 | 3.8 (5.2) | 1.00 (0.85-1.16) | 0.185 |
|  | **2019** | 3.9 (4.2) | 1 |  | 4.1 (4.9) | 1 |  | 3.9 (5.0) | 1 |  |
|  | **2020** | 3.4 (4.6) | 0.87 (0.67-1.13) |  | 3.0 (4.6) | 0.73 (0.53-0.99) |  | 3.3 (3.8) | 0.85 (0.70-1.04) |  |
| **MCV** | **2018** | 2.5 (4.2) | 1.01 (0.69-1.46) | 0.421 | 2.5 (3.9) | 0.79 (0.63-1.00) | 0.470 | 2.8 (4.1) | 0.76 (0.57-1.01) | 0.415 |
|  | **2019** | 2.5 (3.4) | 1 |  | 3.1 (4.1) | 1 |  | 3.7 (4.3) | 1 |  |
|  | **2020** | 3.0 (3.6) | 1.18 (0.89-1.56) |  | 2.8 (4.0) | 0.90 (0.61-1.30) |  | 3.1 (4.3) | 0.84 (0.66-1.07) |  |
| **ANC** | **2018** | 5.2 (4.9) | 1.29 (0.97-1.72) | 0.070 | 4.3 (3.8) | 1.11 (0.88-1.40) | 0.149 | 4.4 (3.4) | 1.10 (0.91-1.32) | 0.568 |
|  | **2019** | 4.0 (3.4) | 1 |  | 3.9 (3.5) | 1 |  | 4.0 (4.6) | 1 |  |
|  | **2020** | 4.0 (3.5) | 1.00 (0.85-1.18) |  | 3.6 (3.5) | 0.93 (0.83-1.05) |  | 4.2 (4.0) | 1.03 (0.88-1.22) |  |
| **TT** | **2018** | 5.2 (4.9) | 1.29 (0.97-1.72) | 0.065 | 4.3 (3.8) | 1.11 (0.88-1.40) | 0.149 | 4.5 (3.4) | 1.10 (0.91-1.33) | 0.556 |
|  | **2019** | 4.0 (3.4) | 1 |  | 3.9 (3.5) | 1 |  | 4.0 (4.6) | 1 |  |
|  | **2020** | 4.0 (3.5) | 1.00 (0.85-1.17) |  | 3.6 (3.5) | 0.93 (0.83-1.05) |  | 4.2 (4.0) | 1.03 (0.88-1.22) |  |
| **FP** | **2018** | 4.3 (5.1) | 0.86 (0.70-1.04) | 0.067 | 4.4 (7.4) | 0.82 (0.64-1.06) | 0.007 | 4.1 (4.9) | 0.69 (0.58-0.81) | <0.001 |
|  | **2019** | 5.0 (7.0) | 1 |  | 5.4 (10.3) | 1 |  | 6.0 (10.2) | 1 |  |
|  | **2020** | 5.2 (6.6) | 1.04 (0.86-1.23) |  | 6.5 (11.5) | 1.21 (0.90-1.61) |  | 6.3 (9.7) | 1.06 (0.86-1.30) |  |
| **ART visits (monthly)** | **2018** | 131.6 (102.2) | 1.06 (0.87-1.28) | 0.040 | 132.6 (103.5) | 1.08 (0.87-1.34) | 0.018 | 136.2 (116.2) | 0.89 (0.81-0.97) | 0.288 |
|  | **2019** | 124.6 (83.8) | 1 |  | 122.9 (79.6) | 1 |  | 154.3 (118.0) | 1 |  |
|  | **2020** | 166.1 (153.3) | 1.33 (0.97-1.83) |  | 169.4 (145.2) | 1.38 (1.15-1.64) |  | 172.6 (178.8) | 1.10 (0.76-1.60) |  |
| **New HIV** | **2018** | 0.9 (1.7) | 0.61 (0.30-1.27) | 0.233 | 0.9 (1.5) | 0.66 (0.43-0.99) | 0.080 | 1.1 (1.6) | 0.77 (0.46-1.30) | 0.614 |
|  | **2019** | 1.4 (1.8) | 1 |  | 1.3 (2.0) | 1 |  | 1.4 (1.8) | 1 |  |
|  | **2020** | 1.7 (1.9) | 1.23 (0.63-2.36) |  | 1.1 (1.4) | 0.81 (0.54-1.22) |  | 1.2 (1.5) | 0.85 (0.67-1.10) |  |

**Masaka, Uganda**

Supplementary Figure 5. **Data Availability for each of the main outcomes, the number of facilities with data per week – Masaka, Uganda**

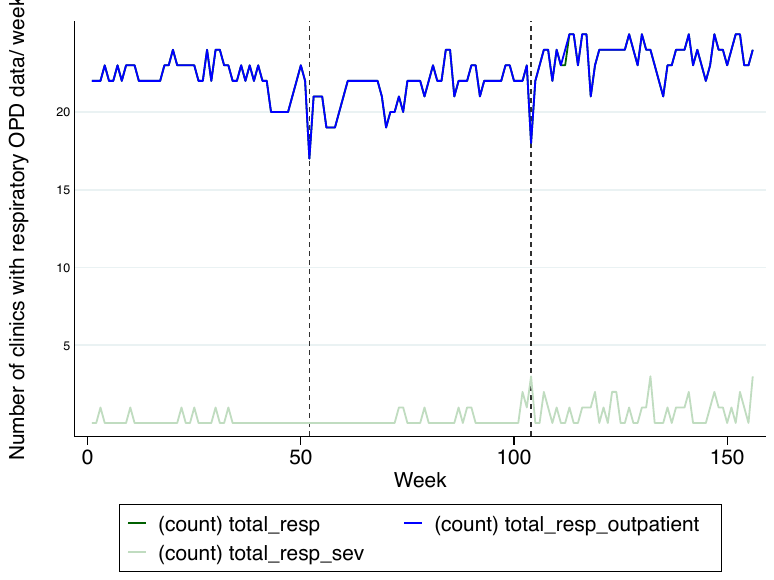

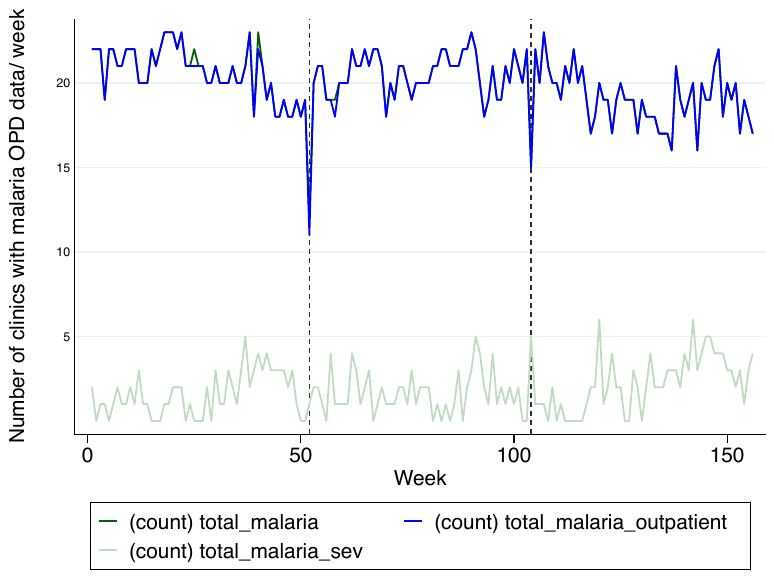

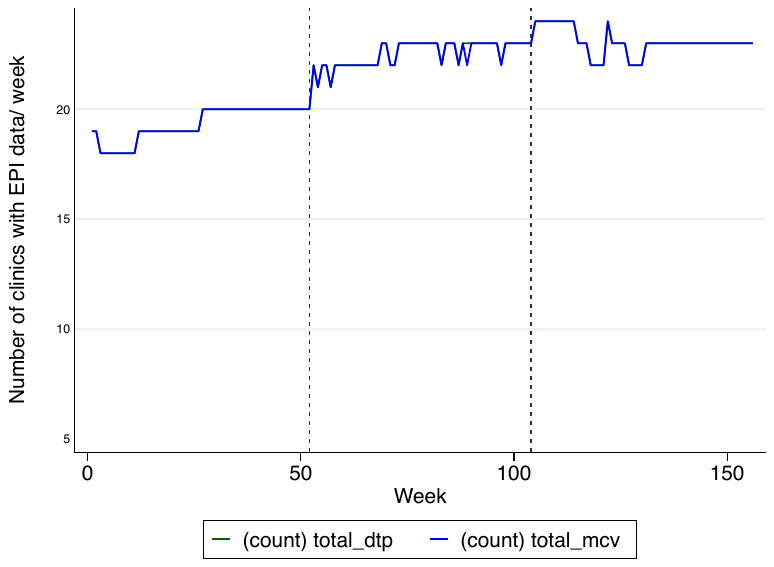

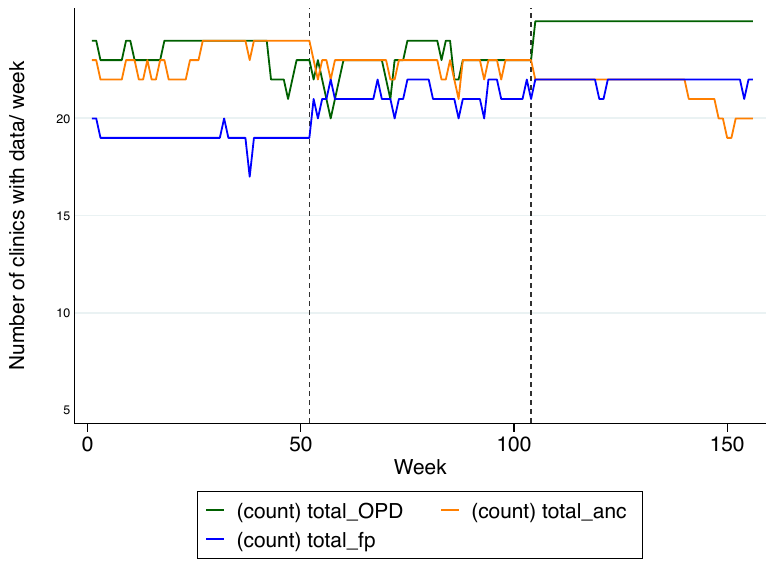

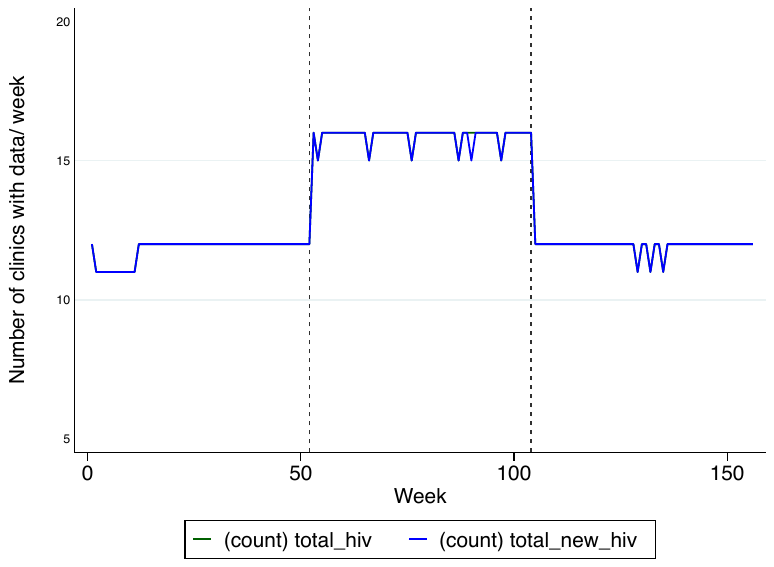

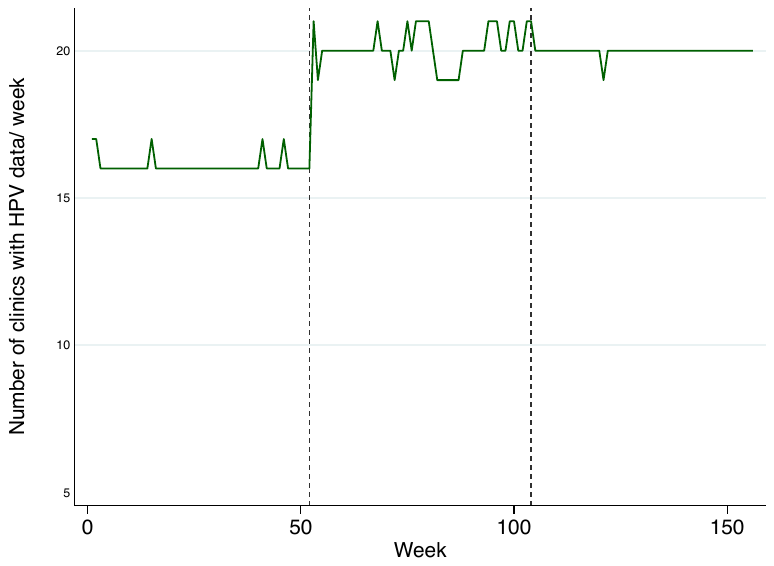

*missing data for severity, and outpatient data for resp/malaria

Abbreviations: ANC: Ante-natal care; ART: Anti-retroviral treatment; DTP: Diphtheria-tetanus-pertussis vaccine (3^rd^ dose)/ pentavalent vaccine (third dose); FP: Family Planning; HIV: Human Immunodeficiency Virus; MCV: Measles containing vaccine (1^st^ dose); OPD: under-5 outpatient department visit; Resp: respiratory outpatient department visits; TT: Tetanus toxoid vaccination

Supplementary Figure 6. **Plots of the mean number of consultations per facility, per week for specific services, indicted on the y axis, across each calendar year of the study in 25 facilities in Masaka, Uganda.** Vertical grey lines are the periods of analysis.

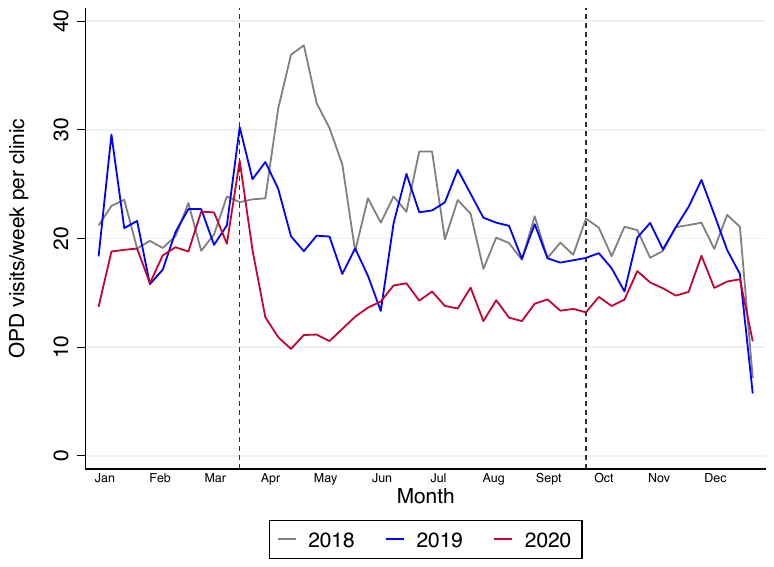

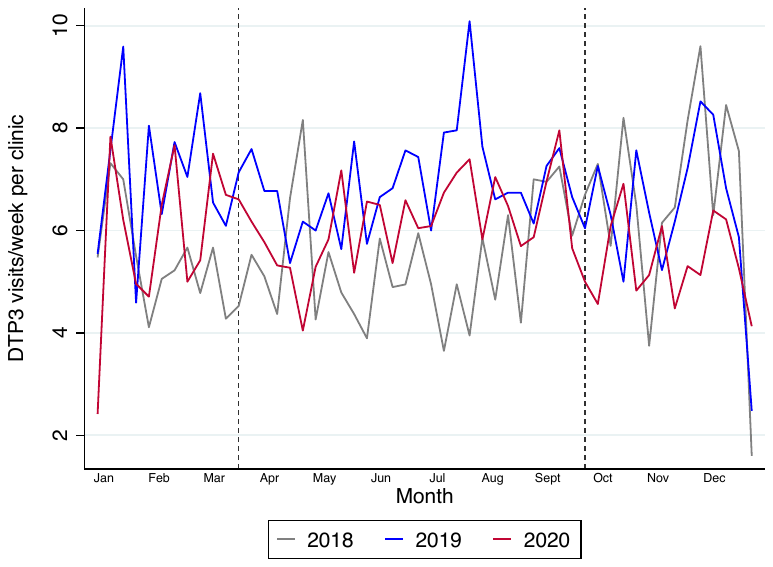

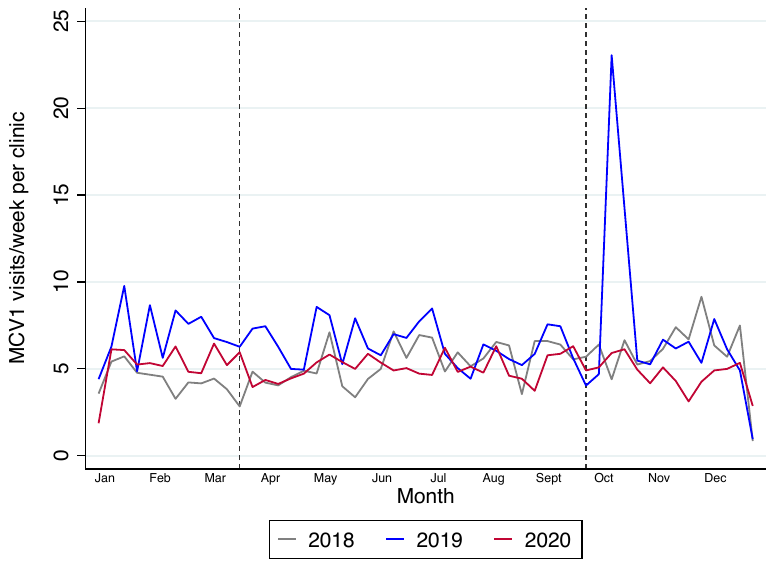

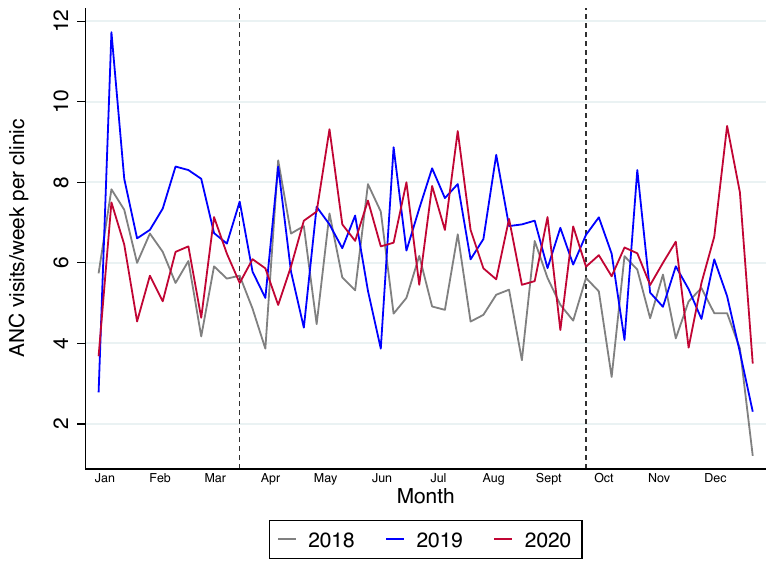

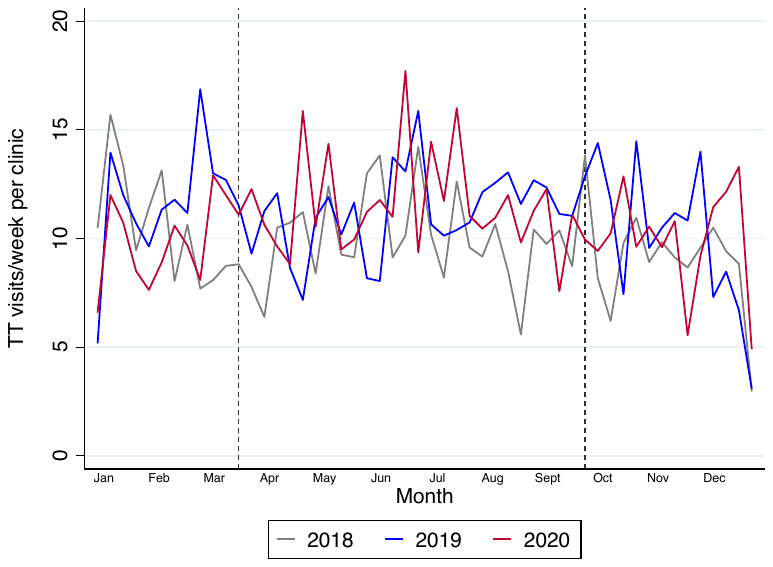

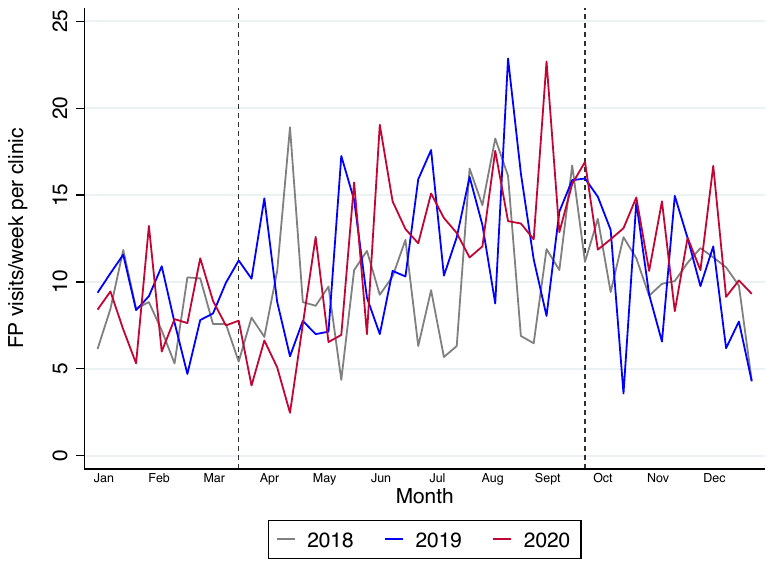

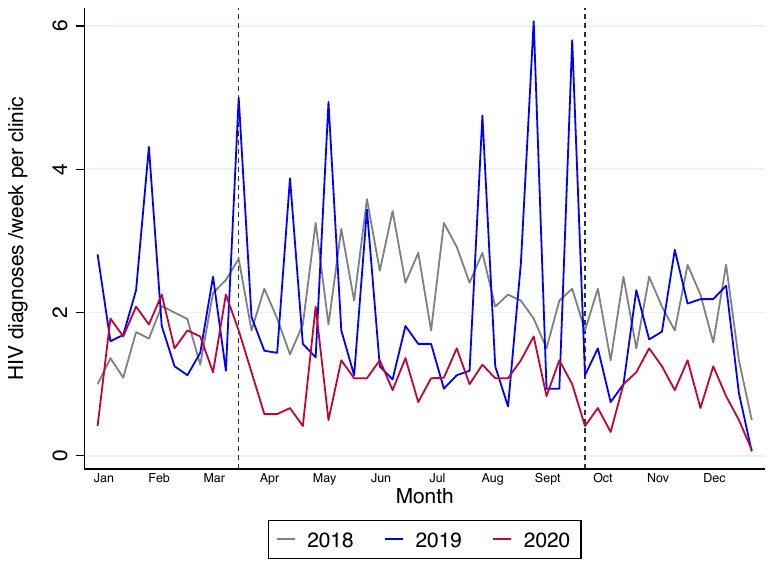

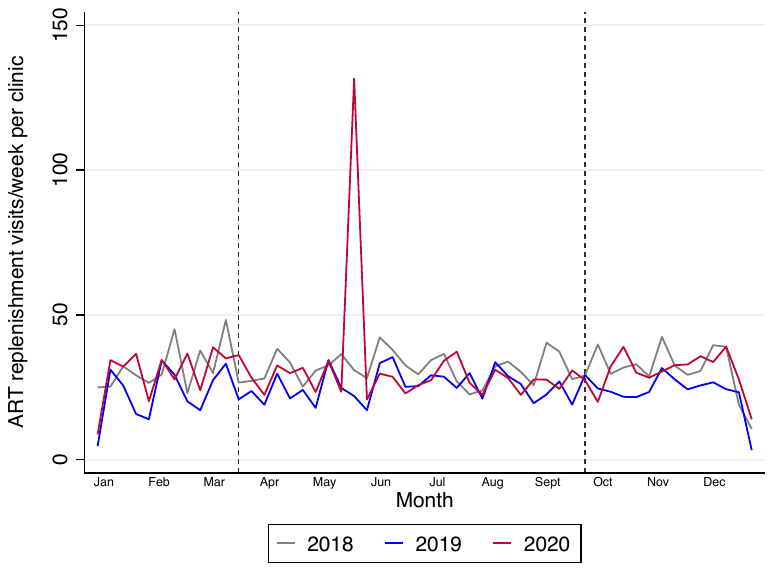

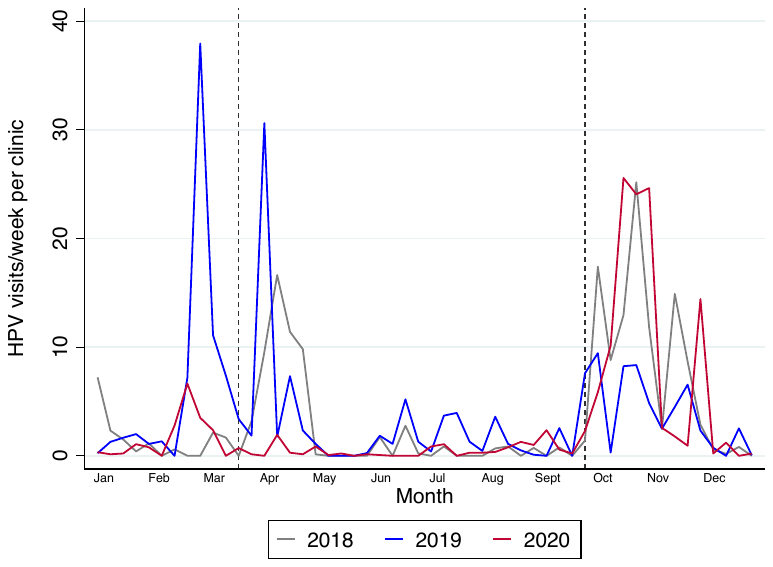

Supplementary Table 3: **Comparison of mean activity levels in similar calendar periods in 2018, 2019 and 2020 using a negative binomial regression model accounting for clustering by facility, and controlling for month (season) – in Masaka, Uganda**

|  |  | **01Jan-15Mar 2020**  **(pre-epidemic)** | | | **16 March – 20^th^ Sept 2020**  **Period 1 of lockdown** | | | **21 Sept – 27^th^ Dec 2020**  **Period 2 of lockdown** | | |
| --- | --- | --- | --- | --- | --- | --- | --- | --- | --- | --- |
| **Outcome** | **Year** | **Mean visits/week (s.d)** | **Mean ratio** | **p-value^2^** | **Mean visits/week (s.d)** | **Mean ratio** | **p-value^2^** | **Mean visits/week (s.d)** | **Mean ratio** | **p-value^2^** |
| **OPD visits** | **2018** | 21.1 (14.3) | 1.01 (0.82-1.24) | 0.135 | 24.1 (17.5) | 1.12 (0.98-1.28) | <0.001 | 19.5 (12.9) | 1.04 (0.94-1.15) | 0.010 |
|  | **2019** | 21.0 (12.8) | 1 |  | 21.4 (14.8) | 1 |  | 18.8 (13.9) | 1 |  |
|  | **2020** | 18.8 (13.2) | 0.90 (0.74-1.09) |  | 13.9 (10.3) | 0.65 (0.58-0.73) |  | 15.1 (10.7) | 0.80 (0.67-0.97) |  |
| **Resp OPD** | **2018** | 12.2 (7.1) | 0.90 (0.77-1.05) | 0.401 | 13.7 (9.4) | 1.12 (0.99-1.26) | <0.001 | 11.6 (6.8) | 1.19 (1.06-1.34) | <0.001 |
|  | **2019** | 13.5 (8.0) | 1 |  | 12.2 (8.3) | 1 |  | 9.8 (6.9) | 1 |  |
|  | **2020** | 11.6 (8.1) | 0.85 (0.70-1.03) |  | 7.7 (6.4) | 0.64 (0.55-0.74) |  | 7.9 (5.3) | 0.81 (0.67-0.97) |  |
| **Malaria OPD** | **2018** | 3.5 (2.8) | 0.98 (0.81-1.18) | 0.037 | 4.7 (4.3) | 0.95 (0.75-1.18) | <0.001 | 2.8 (3.0) | 0.79 (0.64-0.97) | 0.073 |
|  | **2019** | 3.6 (2.9) | 1 |  | 4.8 (4.2) | 1 |  | 3.6 (3.3) | 1 |  |
|  | **2020** | 4.3 (3.3) | 1.21 (0.96-1.53) |  | 2.7 (2.3) | 0.54 (0.46-0.64) |  | 2.3 (2.2) | 0.63 (0.52-0.76) |  |
| **DTP** | **2018** | 5.5 (4.7) | 0.78 (0.51-1.18) | 0.629 | 5.4 (5.0) | 0.77 (0.58-1.02) | 0.391 | 6.6 (7.1) | 1.04 (0.87-1.23) | 0.077 |
|  | **2019** | 7.1 (11.7) | 1 |  | 6.9 (7.6) | 1 |  | 6.4 (8.1) | 1 |  |
|  | **2020** | 5.9 (6.1) | 0.83 (0.64-1.08) |  | 6.2 (6.5) | 0.89 (0.77-1.02) |  | 5.4 (5.7) | 0.85 (0.69-1.04) |  |
| **MCV** | **2018** | 4.4 (4.5) | 0.63 (0.41-0.99) | 0.318 | 5.3 (5.0) | 0.82 (0.62-1.09) | 0.776 | 6.0 (7.2) | 0.44 (0.15-1.28) | 0.011 |
|  | **2019** | 7.0 (12.0) | 1 |  | 6.4 (8.0) | 1 |  | 15.2 (149) | 1 |  |
|  | **2020** | 5.2 (5.2) | 0.75 (0.53-1.05) |  | 5.1 (5.3) | 0.79 (0.68-0.91) |  | 4.7 (5.3) | 0.34 (0.12-0.95) |  |
| **ANC** | **2018** | 6.1 (5.3) | 0.83 (0.68-1.00) | 0.652 | 5.6 (5.4) | 0.83 (0.75-0.93) | 0.081 | 4.7 (4.7) | 0.87 (0.72-1.04) | 0.072 |
|  | **2019** | 7.4 (9.0) | 1 |  | 6.7 (6.6) | 1 |  | 5.4 (5.7) | 1 |  |
|  | **2020** | 5.8 (5.0) | 0.78 (0.56-1.09) |  | 6.6 (6.8) | 0.98 (0.79-1.21) |  | 6.1 (8.4) | 1.14 (0.79-1.66) |  |
| **TT** | **2018** | 10.6 (10.8) | 0.91 (0.81-1.02) | 0.428 | 9.9 (11.5) | 0.89 (0.79-1.00) | 0.071 | 9.1 (10.0) | 0.90 (0.77-1.06) | 0.490 |
|  | **2019** | 11.7 (11.4) | 1 |  | 11.2 (11.0) | 1 |  | 10.2 (11.7) | 1 |  |
|  | **2020** | 9.8 (10.3) | 0.84 (0.68-1.03) |  | 11.6 (14.2) | 1.04 (0.88-1.22) |  | 10.0 (14.8) | 1.00 (0.72-1.39) |  |
| **FP** | **2018** | 8.3 (8.4) | 0.93 (0.73-1.19) | 0.937 | 10.4 (18.3) | 0.87 (0.64-1.17) | 0.589 | 10.5 (10.7) | 1.03 (0.78-1.35) | 0.385 |
|  | **2019** | 8.9 (9.4) | 1 |  | 12.0 (21.2) | 1 |  | 10.4 (16.3) | 1 |  |
|  | **2020** | 8.5 (13.3) | 0.95 (0.66-1.36) |  | 11.7 (15.7) | 0.94 (0.69-1.29) |  | 12.2 (15.1) | 1.20 (0.90-1.61) |  |
| **ART visits (monthly)** | **2018** | 31.9 (36.3) | 1.41 (1.02-1.94) | 0.548 | 31.6 (35.3) | 1.24 (0.92-1.69) | 0.943 | 31.1 (34.0) | 1.32 (0.93-1.87) | 0.830 |
|  | **2019** | 22.9 (33.9) | 1 |  | 25.4 (34.5) | 1 |  | 23.6 (30.7) | 1 |  |
|  | **2020** | 29.9 (31.9) | 1.32 (0.95-1.84) |  | 32.0 (77.1) | 1.26 (0.83-1.90) |  | 30.3 (29.2) | 1.29 (0.93-1.78) |  |
| **New HIV** | **2018** | 1.7 (1.9) | 0.85 (0.45-1.60) | 0.908 | 2.4 (2.4) | 1.06 (0.70-1.60) | <0.001 | 1.9 (2.5) | 1.18 (0.76-1.81) | <0.001 |
|  | **2019** | 2.0 (3.9) | 1 |  | 2.3 (6.8) | 1 |  | 1.6 (2.3) | 1 |  |
|  | **2020** | 1.7 (1.9) | 0.84 (0.43-1.63) |  | 1.1 (1.8) | 0.49 (0.25-0.95) |  | 0.9 (1.2) | 0.52 (0.32-0.87) |  |
| **HPV** | **2018** | 1.5 (11.4) | 0.29 (0.05-1.60) | 0.924 | 2.2 (11.7) | 0.67 (0.31-1.47) | 0.008 | 7.7 (23.9) | 1.78 (0.95 – 3.35) | 0.909 |
|  | **2019** | 6.5 (39.9) | 1 |  | 2.9 (14.8) | 1 |  | 4.2 (14.3) | 1 |  |
|  | **2020** | 1.6 (6.0) | 0.27 (0.06-1.24) |  | 0.5 (2.2) | 0.23 (0.09-0.57) |  | 8.3 (20.0) | 1.84 (0.91-3.72) |  |

^2^ The document p-values are Wald tests, testing the hypothesis that the coefficients for ‘year’ estimated by the negative binomial regression model, are equal. Likelihood ratio tests are invalid given that the likelihood estimated under robust standard errors does not account for clustering: <https://www.stata.com/support/faqs/statistics/likelihood-ratio-test/>

**Supplementary material: Climate data from WeatherSpark.com**

Lungi International Airport, Freetown, Sierra Leone

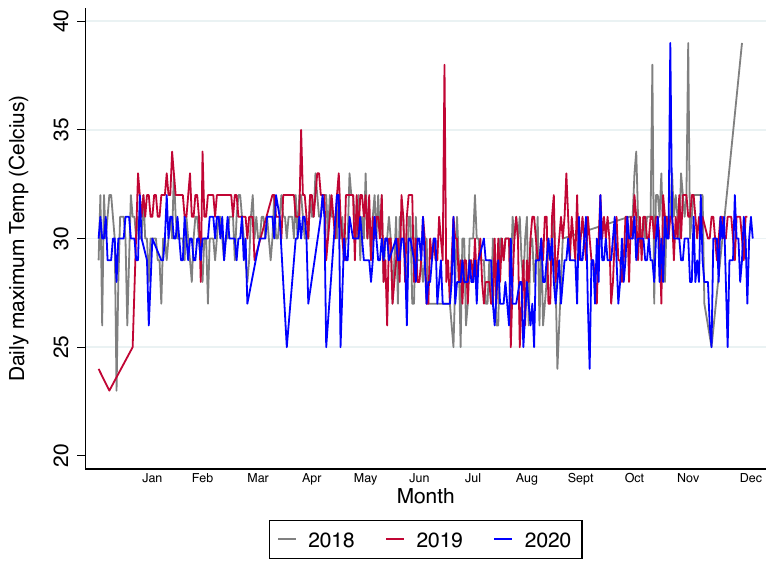

Entebbe International Airport, Uganda

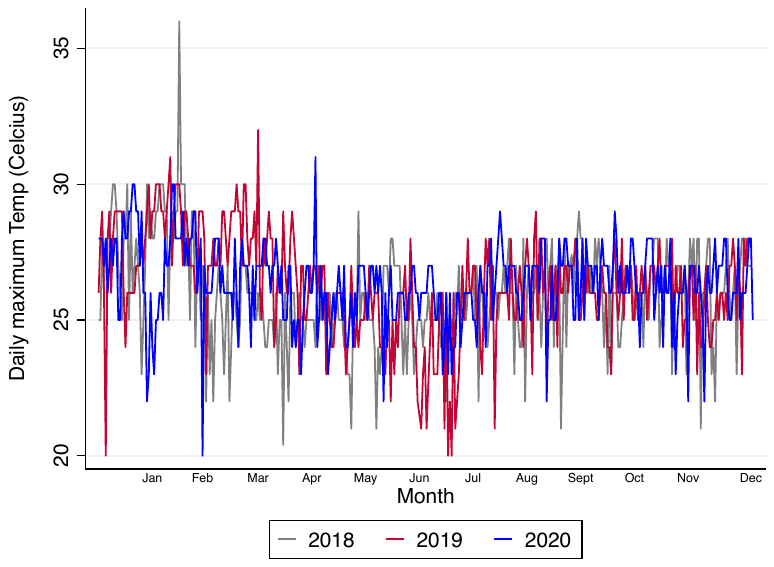
